# Circulating-blood dose is associated with severe lymphopenia in thoracic radiotherapy, but added simulation complexity provides no detectable incremental information

**DOI:** 10.64898/2026.08.13.26359883

**Authors:** Weisi Yan, Yajing Wu, Xuan Liang, Adia Holtman, James Castle, Donlin Yan, Mingming Ge, Suyang Zou, Yao Zhang, Shufan Yue, Thomas Oldland, Ron McGarry, Ellis Johnson, Dennis Cheek, Jun Wang

**Affiliations:** Department of Radiation Oncology, the Fourth Hospital of Hebei Medical University, Hebei Clinical Research Center for Radiation Oncology; Department of Radiation Medicine, Markey Cancer Center, University of Kentucky – College of Medicine, USA

**Author notes:** Co-corresponding authors: Weisi Yan M.D.,Ph.D., University of Kentucky College of Medicine, Radiation Medicine, 800 Rose St., C111 Pav H Lexington, KY 40536-0086, Jun Wang, MD, Department of Radiation Oncology, the Fourth Hospital of Hebei Medical University, Hebei Clinical Research Center for Radiation Oncology, No. 12 Jiankang Road, Shijiazhuang, Hebei 050011, China. Weisi Yan and Yajing Wu contributed equally to this work.

**Keywords:** ICE3, radiation-induced lymphopenia, circulating blood dose, EDIC, HEDOS, planning target volume, thoracic radiation therapy

## Abstract

**Purpose:** To compare scalar circulating-blood dose summaries, assess whether associations with grade 3 or higher lymphopenia and overall survival persisted after planning target volume (PTV) adjustment, and distinguish scalar from exploratory dynamic blood-dose analyses.

**Methods and Materials:** The assembled dataset included 133 patients from 2 retrospective thoracic radiation cohorts; 121 entered the dosimetric main analysis and 116 passed a post hoc dosimetric gate. Six scalar exposures were compared: an ICE3 (Immune Circulation radiation Exposure Estimator Engine) 10-compartment mean-dose metric, corrected effective dose to immune cells (EDIC), body-remainder dose, a hematological dose (HEDOS)-derived organ-mean approximation, mean lung dose, and mean heart dose. Logistic base models included baseline absolute lymphocyte count, concurrent chemotherapy, and cohort; PTV was then added. Overall survival used cohort-stratified Cox models on an endpoint-specific common set. Benjamini-Hochberg correction was applied within prespecified 6-exposure families and, separately, across 3 selected post hoc bootstrap contrasts.

**Results:** The lymphopenia analysis included 94 patients and 69 events. After PTV adjustment, the HEDOS-derived approximation remained nominally associated (odds ratio, 2.54; 95% confidence interval, 1.18-5.45; P=.017; q=.102), but no exposure survived false-discovery-rate control. The survival set contained 92 patients and 42 deaths; no PTV-adjusted scalar exposure was associated with survival (all q>=.531). In post hoc analyses, the standardized association of mean heart dose with overall survival was more positive than that of ICE3 (difference in log hazard ratios, 0.44; 95% CI, 0.13–0.94; multiplicity-adjusted q=.024). The corresponding contrast with the HEDOS-derived approximation did not meet the adjusted significance threshold (q=.053). These coefficient contrasts do not establish superior predictive performance or causality. Exported ICE3 kinetic summaries had no false-discovery-rate-significant residual associations. A 13-case HEDOS bDVH audit showed little change under one continuous-versus-10-second-gap perturbation.

**Conclusions:** PTV adjustment attenuated scalar blood-dose associations with severe lymphopenia, and no scalar exposure retained a multiplicity-robust survival association. The selected mean-heart- dose coefficient contrast is hypothesis-generating and does not establish superior prediction. The primary cohort comparison evaluated a HEDOS-derived organ-mean approximation rather than the full dynamic HEDOS framework; therefore, these findings should not be interpreted as evidence against the potential value of particle-level blood-dose distributions or time-dependent blood-flow modeling.

## Introduction

Radiation-induced lymphopenia is common during thoracic radiation therapy and has been associated with adverse outcomes in several disease settings. The biological rationale is plausible: circulating lymphocytes are radiosensitive, thoracic radiation fields expose vascular and lymphoid compartments, and systemic therapy can contribute to hematologic toxicity.^1–5^

Several approaches estimate radiation exposure to circulating immune cells: EDIC and multi-compartment methods produce scalar summaries from organ or compartment doses, whereas HEDOS models stochastic blood movement and time-dependent radiation delivery to generate blood-dose distributions.^6–13^ Even when ICE3 mean dose or another scalar metric can be reduced to a weighted combination of organ means, this does not make a dynamic HEDOS-type model redundant. A dynamic model may still provide clinically relevant distributional and repeated-exposure information that cannot be recovered from the mean alone.

The primary objective was to compare 6 scalar dose summaries on identical patients and evaluate whether their associations with grade 3 or higher lymphopenia and overall survival remained after conditioning on PTV. The comparison was not designed as an equivalence, noninferiority, or predictive-performance study. Because the full cohorts lacked the complete dosimetric and treatment- delivery inputs required for dynamic blood-flow modeling, dynamic analyses were exploratory. We therefore evaluated previously exported ICE3 kinetic summaries and, in a small subset with suitable DVH and treatment-timing data, assessed the feasibility of generating HEDOS blood dose-volume histograms. These subset analyses were not intended to provide a definitive head-to-head comparison of full dynamic HEDOS with scalar metrics.

## Methods and Materials

### Study Design and Participants

The retrospective assembled dataset contained 133 patients from 2 institutional thoracic radiation cohorts. Cohorts are labeled A and B in the anonymized manuscript. The dosimetric main-analysis cohort contained 121 patients; 116 passed the post hoc dosimetric gate. The gated set included 80 lung primaries, 33 esophageal primaries, and 3 other or unknown primaries. Cohort A contained 33 lung and 33 esophageal primaries. Cohort B contained 47 lung and 3 other or unknown primaries, including 22 small-cell lung cancers.

Complete-case analysis was used for each endpoint; no imputation was performed. The primary lymphopenia common set contained 94 patients with all 6 exposures, baseline absolute lymphocyte count (ALC), concurrent-chemotherapy status, cohort, PTV, and lymphopenia outcome available. The endpoint-specific survival common set contained 92 patients with all 6 exposures, PTV, cohort, survival time, and event status available.

This retrospective study was approved by the institutional review boards of the participating institutions with a waiver of informed consent for use of existing clinical, laboratory, and radiotherapy data. For this anonymized manuscript, institutions are labeled Cohort A and Cohort B and protocol identifiers are withheld. This report follows the Strengthening the Reporting of Observational Studies in Epidemiology (STROBE) reporting guideline for observational studies.^14^

### Endpoints

The primary endpoint was grade 3 or higher lymphopenia based on the ALC nadir from treatment initiation through 30 days after radiation completion. Overall survival was the time-to-event endpoint. Progression-free survival was examined only in a post hoc dynamic audit.

Lymphopenia was graded according to CTCAE version 5.0 using absolute lymphocyte count in x10^9/L: grade 0, >=1.0; grade 1, 0.8 to <1.0; grade 2, 0.5 to <0.8; grade 3, 0.2 to <0.5; and grade 4, <0.2. Grade 3 or higher lymphopenia was therefore defined as ALC nadir <0.5 x10^9/L. Baseline ALC was the last ALC value before or on the radiotherapy start date. The primary ALC nadir window was radiotherapy start through 30 days after radiotherapy completion.

Overall survival was measured from radiotherapy start to death from any cause and was censored at the last known alive or last-contact date in the source records. Cohort A survival events were independently adjudicated from the source workbooks; repeated dates such as April 2, 2026, were treated as follow-up/censoring dates when the event flag and event-site fields did not support a true event. Cohort B used the source OS days and OS event fields from the clinical extraction.

Progression-free survival was post hoc. In the locked derivation, PFS time was measured from radiotherapy start to the source-recorded progression/censoring time. Cohort A used the adjudicated progression indicator and Cohort B used the source PFS event field. Death without recorded progression was not independently added to the Cohort A progression-free-survival event indicator in the locked frame. This derivation therefore departs from the conventional definition of progression-free survival as progression or death from any cause, and the post hoc progression-free-survival results should be read with that limitation in mind. Cohort A records contain follow-up and event dates through April 2, 2026; the Cohort B source frame has populated death dates through January 29, 2026 and populated progression dates through September 15, 2025.

### Scalar Dose Metrics

Six scalar exposures were compared: a corrected ICE3 10-compartment circulating-blood mean-dose metric, corrected EDIC, body-remainder dose, a HEDOS-derived organ-mean approximation, mean lung dose, and mean heart dose. Effects were expressed per standard deviation within the relevant common analysis set.

For the full cohort, the ICE3 exposure was a single whole-course mean blood-dose estimate calculated as a weighted combination of compartment mean doses. This scalar summarizes average exposure but does not describe the distribution or timing of blood irradiation, such as high-dose exposure, the proportion of blood receiving a specified dose, or repeated irradiation of the same circulating blood cells. Likewise, the full-cohort HEDOS-derived exposure was an organ-mean approximation and should not be interpreted as full dynamic HEDOS.^12,13^

All scalar dose metrics are whole-course doses in Gy. The engine used was ICE3 v2.0 (internal source designation v09_engine/v09_engine_10c). At the time of posting no immutable public release identifier had been deposited for this engine, so the metric is referred to throughout as the ICE3 10- compartment circulating-blood mean-dose metric.

For the ICE3 mean endpoint, the expected value has the closed form D_ICE3 = sum_j pi_j D_j, where D_j is the compartment mean dose and pi_j is the steady-state occupancy weight. The 10 compartments and weights are: body, 52/60 = 0.866667; lungs, 5/60 = 0.083333; right atrium, 0.5/60 = 0.008333; right ventricle, 0.5/60 = 0.008333; pulmonary artery, 0.4/60 = 0.006667; pulmonary veins, 0.3/60 = 0.005000; left atrium, 0.3/60 = 0.005000; left ventricle, 0.4/60 = 0.006667; aorta, 0.5/60 = 0.008333; and coronary branch, 0.05 x 2.0/60 = 0.001667. The cardiac compartments sum to 3/60 = 0.050000.

The corrected body-remainder term was D_body-rem = f_blood x mean dose in BODY excluding PTV, lung, and heart, where f_blood is the ICRP-89 blood-volume fraction represented by the imaged body remainder. The corrected ICE3 metric used in the locked v19 analysis was D_ICE3,corr = D_ICE3,locked - (52/60) x MBD_locked + (52/60) x D_body-rem. In the v19 frame, this identity reproduced the stored corrected ICE3 column within floating-point precision: n=119, slope 1.000000, intercept 8.88e-16 Gy, mean absolute error 5.78e-17 Gy, and maximum absolute error 8.88e-16 Gy. Corrected EDIC used the frozen EDIC structure D_EDIC = 0.12 x MLD + 0.08 x MHD + c_i x MBD, with c_i = 0.45 + 0.2975 x sqrt(n_fractions/45) in the published formula. The implementation reconstructed the per-patient body coefficient from the locked EDIC value as c_i = (D_EDIC,locked - 0.12 x MLD - 0.08 x MHD) / MBD_locked and then applied the same body substitution: D_EDIC,corr = D_EDIC,locked - c_i x MBD_locked + c_i x D_body-rem. This identity reproduced the corrected EDIC column within floating-point precision: n=123, slope 1.000000, intercept −4.44e- 16 Gy, mean absolute error 7.94e-17 Gy, and maximum absolute error 8.88e-16 Gy. The published EDIC formulation normalises the integral total dose to a fixed reference body volume of 61.8 × 10³ cm³ rather than to the patient’s own contoured body volume; the body term used here is therefore an approximation whenever the contoured body volume departs from that reference.^8^

The body-remainder dose metric was the scalar D_body-rem above. In the v19 frame, D_body-rem = f_blood x mean dose in BODY excluding PTV, lung, and heart reproduced the stored body- remainder column within floating-point precision: n=123, slope 1.000000, intercept −2.22e-16 Gy, mean absolute error 3.66e-17 Gy, and maximum absolute error 6.66e-16 Gy.

The full-cohort HEDOS-derived scalar imported into the locked analysis frame was HEDOS_mo_mean_Gy, the mean of a HEDOS multi-organ blood-particle dose distribution generated from organ-group mean doses. The HEDOS run used the ICRP89 phantom, organ groups lung, heart, aorta/large arteries, large veins, stomach/esophagus, red marrow, spongy bone, and compact bone, a 30,000-particle temporal distribution, dt=0.05 s, a 300-s simulation window, start time 5 s, and patient-specific beam-on time capped at 290 s. The main clinical comparison used only the mean scalar, not HEDOS bDVH, repeated-exposure, or upper-tail metrics. The local HEDOS clone is based on MGHPhysicsResearch/hedos commit 139e1a24928e78c5adde313fc1a849c0ffa01e8e.

Closed-form verification used the archived Cohort A engine audit (n=66). The matrix-exponential analytic engine mean regressed on the closed-form occupancy-weighted mean gave slope 0.99999999999999, intercept 6.31e-14 Gy, mean absolute error 7.55e-14 Gy, maximum absolute error 1.85e-13 Gy, and Pearson r=1.000000. The old Pearson r=0.999964 corresponds to the stochastic 20,000-particle Monte Carlo mean versus the closed form, for which slope was 1.0006436631855, intercept 0.011657 Gy, mean absolute error 0.01873 Gy, and maximum absolute error 0.07223 Gy. The analytic engine and the closed form therefore agree to numerical precision; the residual difference against the 20,000-particle Monte Carlo mean reflects expected sampling variation rather than a discrepancy in the underlying model.

### Body-Dose Correction and Dosimetric Gate

Because planning CT scan length varied across patients, the original body-dose term—which averaged dose over the available external contour—was associated with scan length. In the corrected implementation, the body compartment excluded the lungs, heart, and PTV and incorporated in-field scaling to a reference blood volume. Cases were excluded if the lung or heart contours contained no voxels on the dose grid or if the reconstructed mean lung dose was outside 5% to 45% of the prescription dose. Because this quality-control gate was defined after the ungated analysis had been performed, it was not considered outcome-blind. Accordingly, the gated analysis was treated as the primary analysis, with the ungated analysis reported as a sensitivity analysis.

### Exploratory Analyses of Spatial-Temporal Blood-Dose Exposure

To determine whether information beyond the average ICE3 blood dose might be clinically relevant, we examined several ICE3-derived kinetic and distributional variables that were already available in the frozen analysis dataset. These variables were intended to characterize three features not captured by a single mean-dose value: **the range of simulated blood-cell doses**, **the dose accumulated during individual exposure events**, and **the timing and repetition of blood passage through irradiated regions**. Post hoc analyses tested their primary associations and residual associations beyond the ICE3 mean-dose metric for grade 3 or higher lymphopenia, continuous ALC nadir, overall survival, and progression-free survival.

A separate exploratory HEDOS analysis was performed in 13 locally available patients for whom complete organ dose-volume histograms and RTPLAN-derived treatment-timing data were available. For each plan and delivery scenario, the workflow simulated 10,000 blood particles using a 0.05- second time step and generated blood dose-volume histograms under two assumptions: uninterrupted beam delivery and delivery incorporating 10-second gaps between beams. Because plan-level organ DVHs were reused across beams and beam-specific three-dimensional dose-rate distributions were not reconstructed, this analysis should be interpreted as a feasibility and timing-sensitivity assessment rather than a definitive full-cohort comparison of dynamic HEDOS with scalar blood-dose metrics.

### Statistical Analysis

Separate logistic-regression models were fitted for each standardized exposure. The base model included baseline ALC, concurrent chemotherapy, and cohort; the PTV-adjusted model added standardized PTV. Benjamini-Hochberg false-discovery-rate correction was applied across the 6 exposure tests separately within the base and PTV-adjusted families.^15^

Overall survival was analyzed on the endpoint-specific common set containing all 6 scalar exposures. Cox models were stratified by cohort and included the standardized exposure and standardized PTV. A sensitivity set additionally required baseline ALC and chemotherapy. False-discovery-rate correction was applied across the 6 exposure tests. Proportional-hazards diagnostics used scaled Schoenfeld residuals against rank-transformed time.

Three selected PTV-adjusted survival coefficient contrasts were evaluated post hoc by patient-level bootstrap resampling (2000 draws; reported seed, 42). Benjamini-Hochberg correction was applied across these 3 reported contrasts in the present audit. The contrasts compare standardized log hazard-ratio coefficients; they do not directly measure discrimination, calibration, clinical utility, or external predictive performance. No equivalence margin was prespecified, so intervals spanning zero were interpreted as non-detection rather than equivalence.

## Results

### Cohort Composition

The gated main-analysis set contained 116 patients (Table 1). Cohort A was evenly split between lung and esophageal primaries, whereas Cohort B was predominantly lung cancer and included all 22 small-cell lung cancer cases.

**Table 1.**
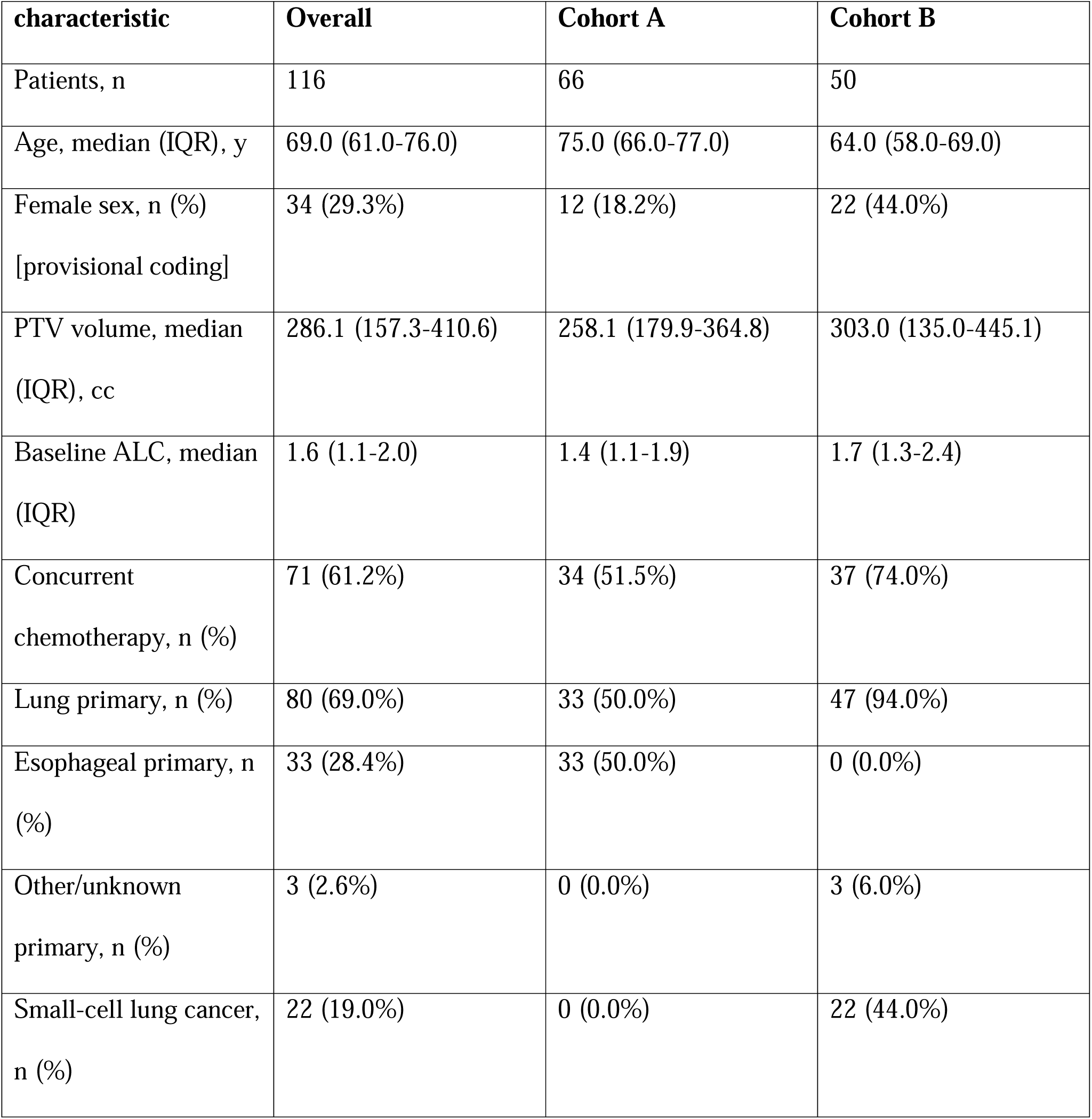
Gated main-analysis set.

### Lymphopenia

Before PTV adjustment, the ICE3 mean-dose metric was associated with grade 3 or higher lymphopenia (odds ratio [OR], 3.07; 95% confidence interval [CI], 1.53-6.17; P=.002; q=.005). After PTV adjustment, ICE3 attenuated to OR, 2.13 (95% CI, 0.99-4.57; P=.053; q=.112), corrected EDIC to OR, 1.98 (95% CI, 0.98-4.00; P=.056; q=.112), and mean lung dose to OR, 1.81 (95% CI, 0.88- 3.75; P=.108; q=.129). The HEDOS-derived organ-mean approximation remained nominally associated (OR, 2.54; 95% CI, 1.18-5.45; P=.017), but not after correction across the 6 PTV-adjusted tests (q=.102). No PTV-adjusted exposure survived false-discovery-rate control (Table 2; Figure 1).

**Figure 1.**
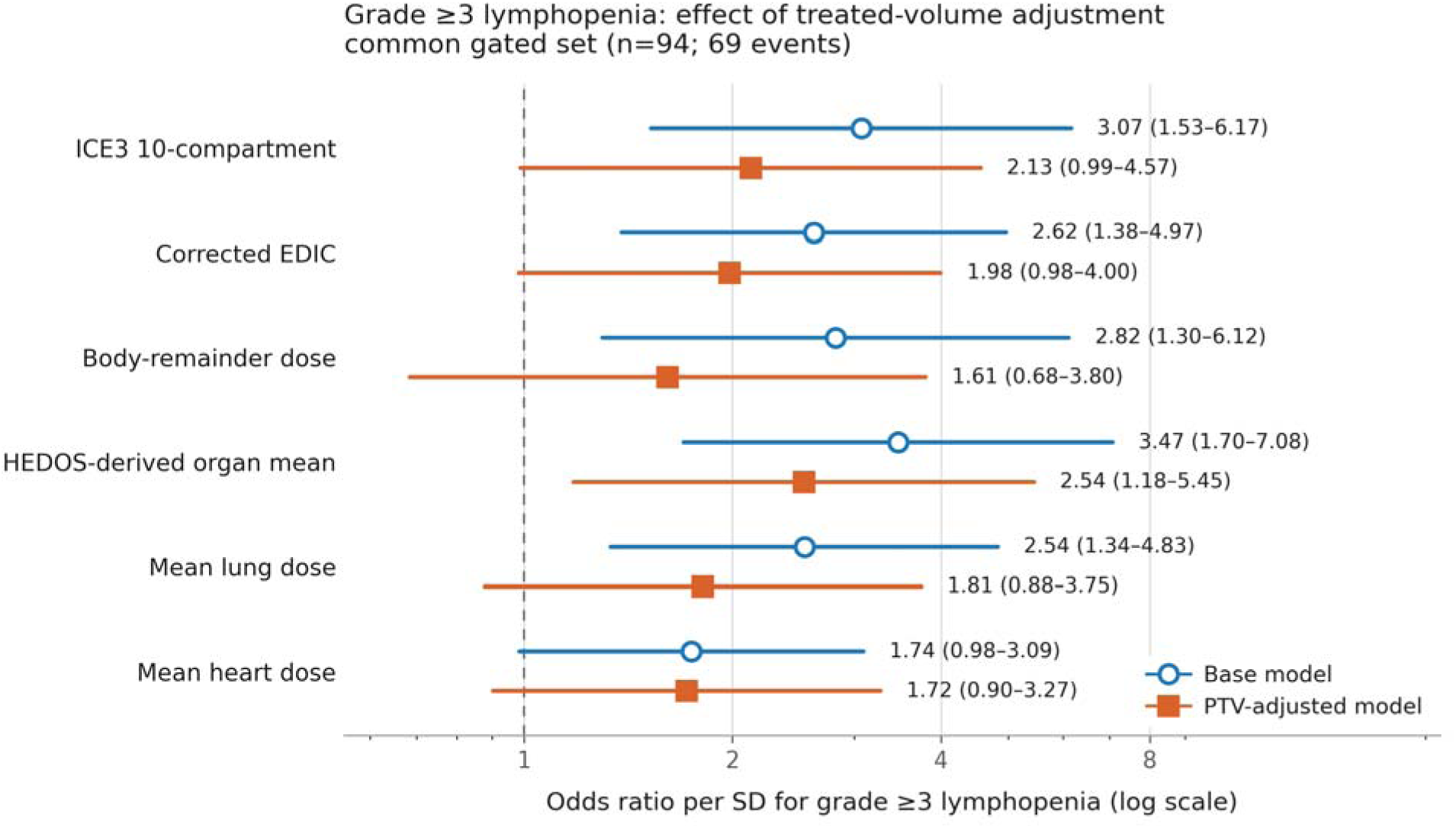
Grade 3 or higher lymphopenia models on the common gated set (n=94; 69 events). Base logistic models included baseline absolute lymphocyte count, concurrent chemotherapy, and cohort. PTV-adjusted models additionally included standardized planning target volume. Points are odds ratios per standard deviation and horizontal bars are 95% confidence intervals. Open circles denote base models and filled squares denote PTV-adjusted models. All 6 scalar exposures are shown. Benjamini-Hochberg q values were calculated within each 6-exposure model family and are reported in Table 2.

**Table 2.**
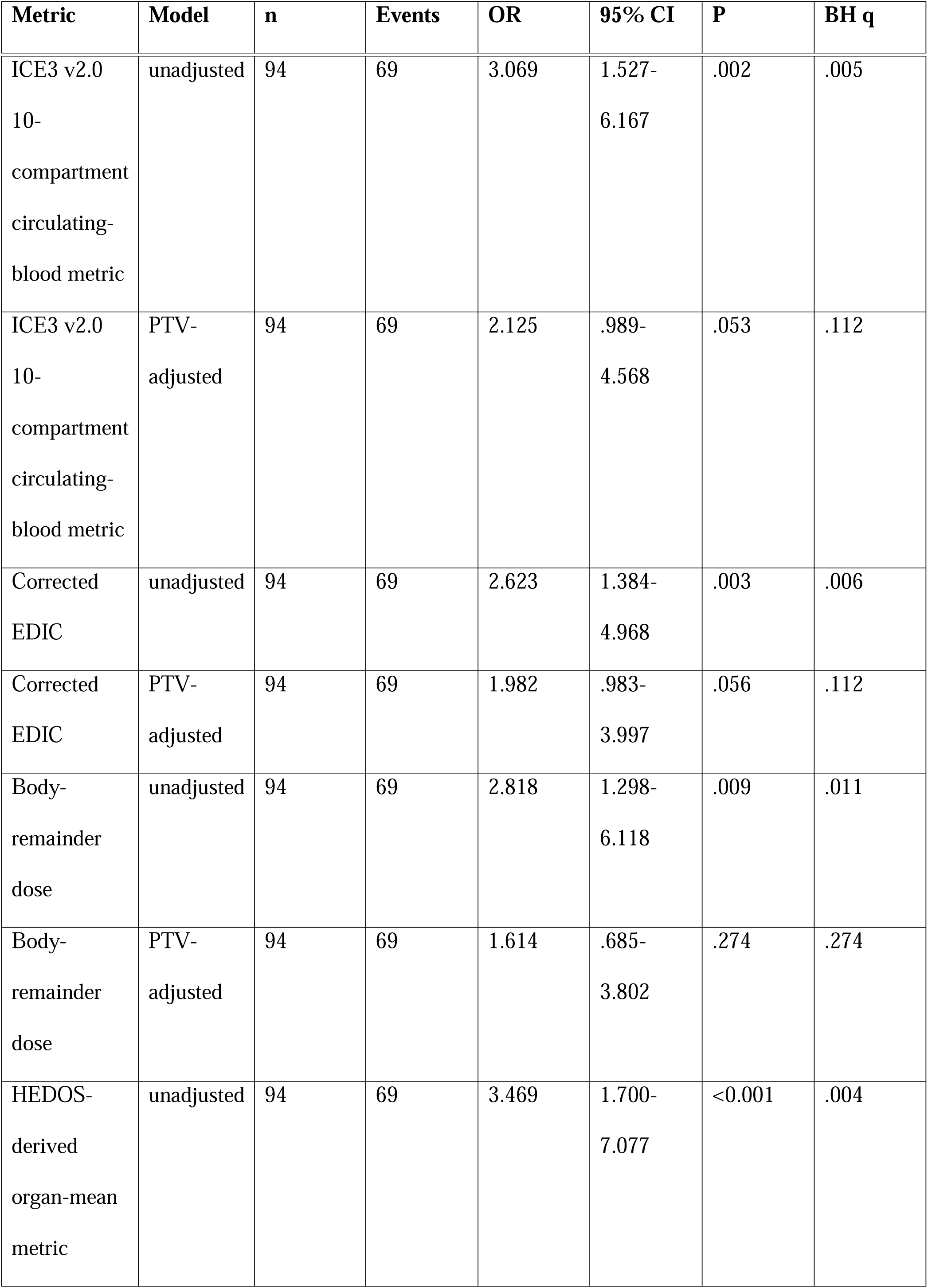
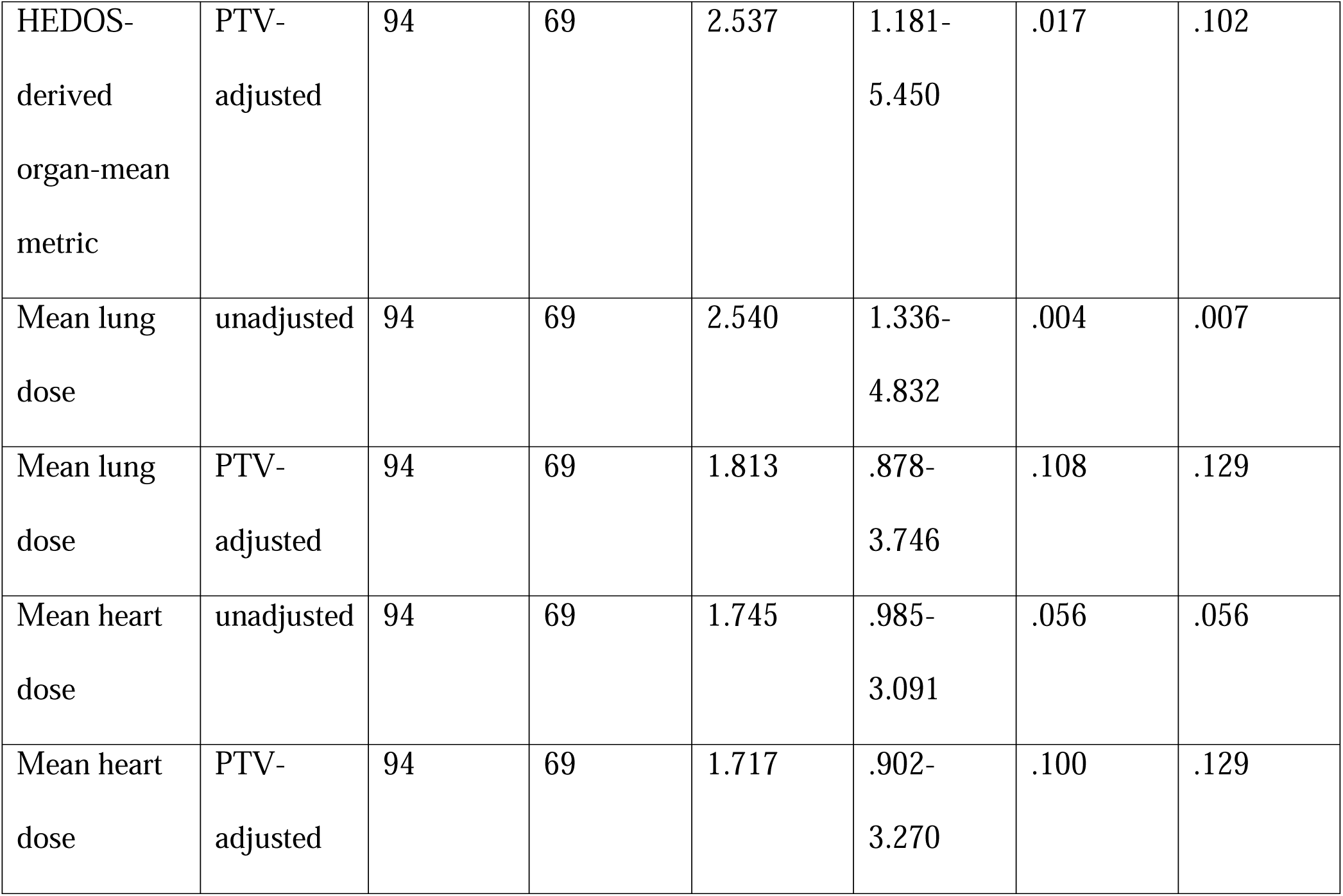
Grade 3 or higher lymphopenia models.

| **Metric** | **Model** | **n** | **Events** | **OR** | **95% CI** | **P** | **BH q** |
| --- | --- | --- | --- | --- | --- | --- | --- |
| ICE3 v2.0<br>10-compartment<br>circulating-blood metric | unadjusted | 94 | 69 | 3.069 | 1.527-6.167 | .002 | .005 |
| ICE3 v2.0<br>10-compartment<br>circulating-blood metric | PTV-adjusted | 94 | 69 | 2.125 | .989-4.568 | .053 | .112 |
| Corrected<br>EDIC | unadjusted | 94 | 69 | 2.623 | 1.384-4.968 | .003 | .006 |
| Corrected<br>EDIC | PTV-adjusted | 94 | 69 | 1.982 | .983-3.997 | .056 | .112 |
| Body-remainder<br>dose | unadjusted | 94 | 69 | 2.818 | 1.298-6.118 | .009 | .011 |
| Body-remainder<br>dose | PTV-adjusted | 94 | 69 | 1.614 | .685-3.802 | .274 | .274 |
| HEDOS-derived<br>organ-mean<br>metric | unadjusted | 94 | 69 | 3.469 | 1.700-7.077 | <0.001 | .004 |
| HEDOS-derived organ-mean metric | PTV-adjusted | 94 | 69 | 2.537 | 1.181-5.450 | .017 | .102 |
| Mean lung dose | unadjusted | 94 | 69 | 2.540 | 1.336-4.832 | .004 | .007 |
| Mean lung dose | PTV-adjusted | 94 | 69 | 1.813 | .878-3.746 | .108 | .129 |
| Mean heart dose | unadjusted | 94 | 69 | 1.745 | .985-3.091 | .056 | .056 |
| Mean heart dose | PTV-adjusted | 94 | 69 | 1.717 | .902-3.270 | .100 | .129 |

### Overall Survival

The endpoint-specific common survival set contained 92 patients and 42 deaths. No PTV-adjusted scalar exposure was associated with overall survival after correction: ICE3, hazard ratio (HR) 0.85 (95% CI, 0.58-1.24; P=.395; q=.592); corrected EDIC, HR 1.09 (95% CI, 0.76-1.57; P=.623; q=.623); body-remainder dose, HR 0.80 (95% CI, 0.51-1.25; P=.318; q=.592); HEDOS-derived approximation, HR 0.90 (95% CI, 0.63-1.30; P=.581; q=.623); mean lung dose, HR 0.82 (95% CI, 0.61-1.12; P=.215; q=.592); and mean heart dose, HR 1.31 (95% CI, 0.96-1.79; P=.089; q=.531) (Table 3; Figure 2).

**Figure 2.**
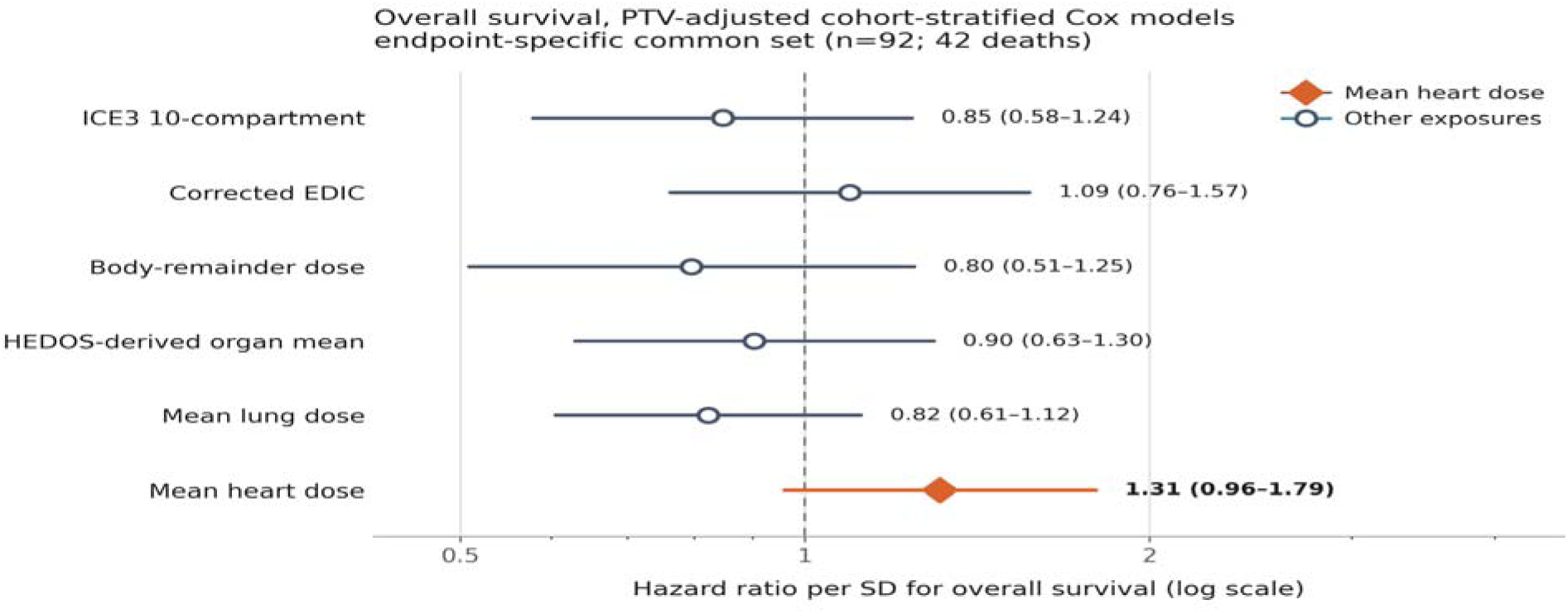
Overall survival on the endpoint-specific common set (n=92; 42 deaths). Cox models were stratified by cohort and included the standardized exposure and standardized planning target volume. Points are hazard ratios per standard deviation and horizontal bars are 95% confidence intervals. The filled diamond identifies mean heart dose; all other exposures are shown as open circles. Benjamini- Hochberg q values were calculated across the 6 exposure tests and are reported in Table 3A.

**Table 3A.**
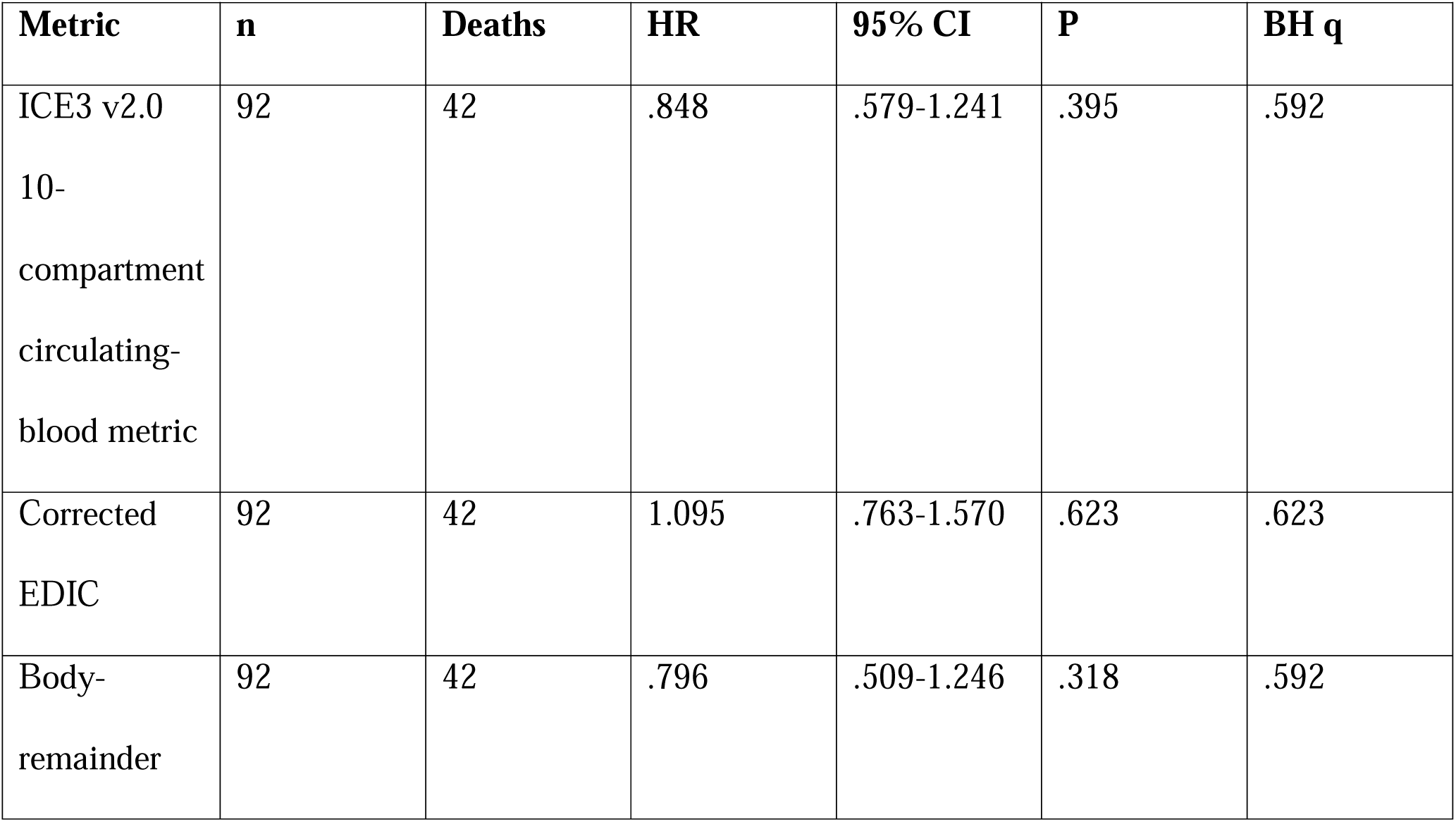
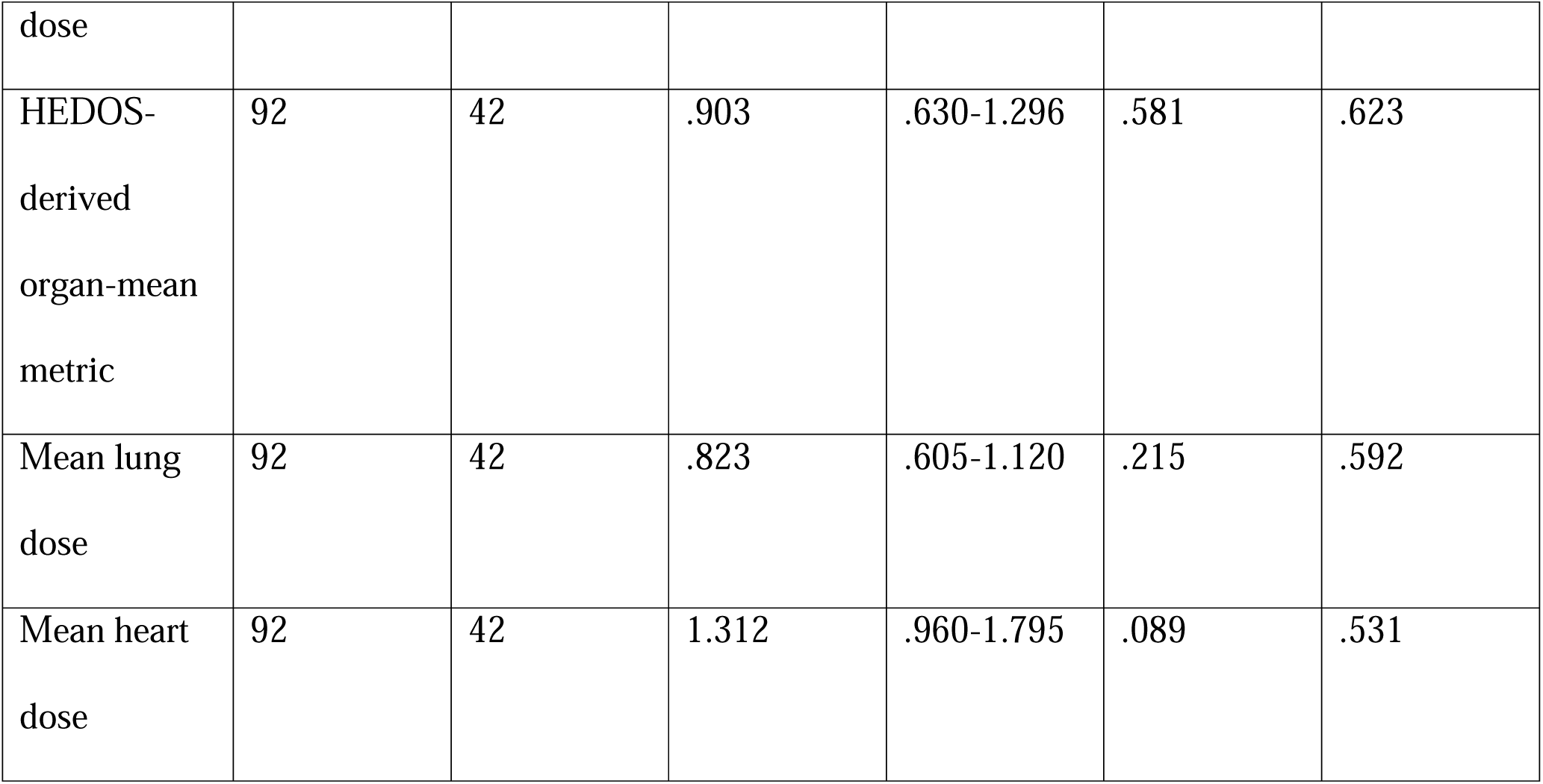
PTV-adjusted Cox models, common survival set.

**Table 3B.**
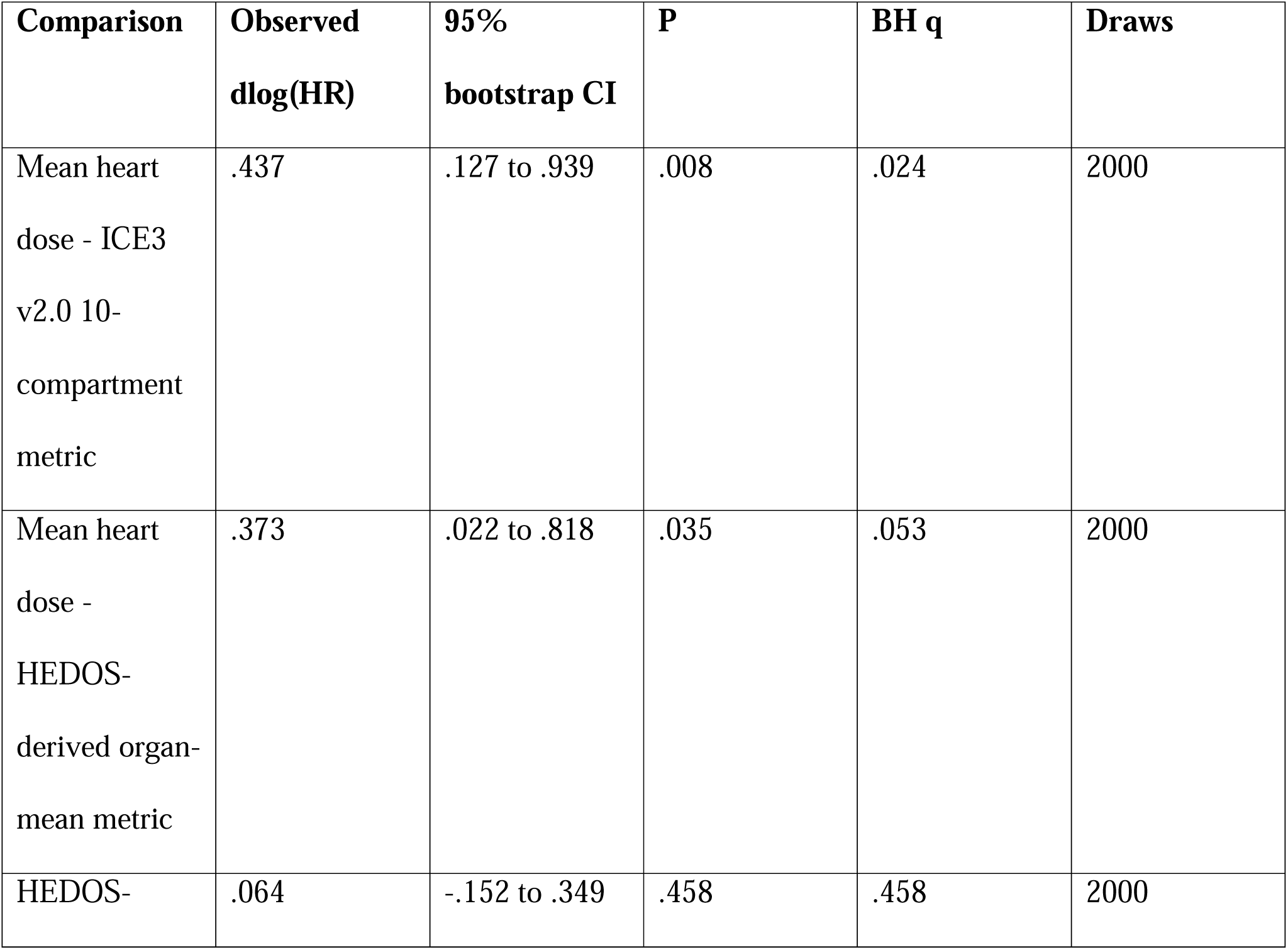
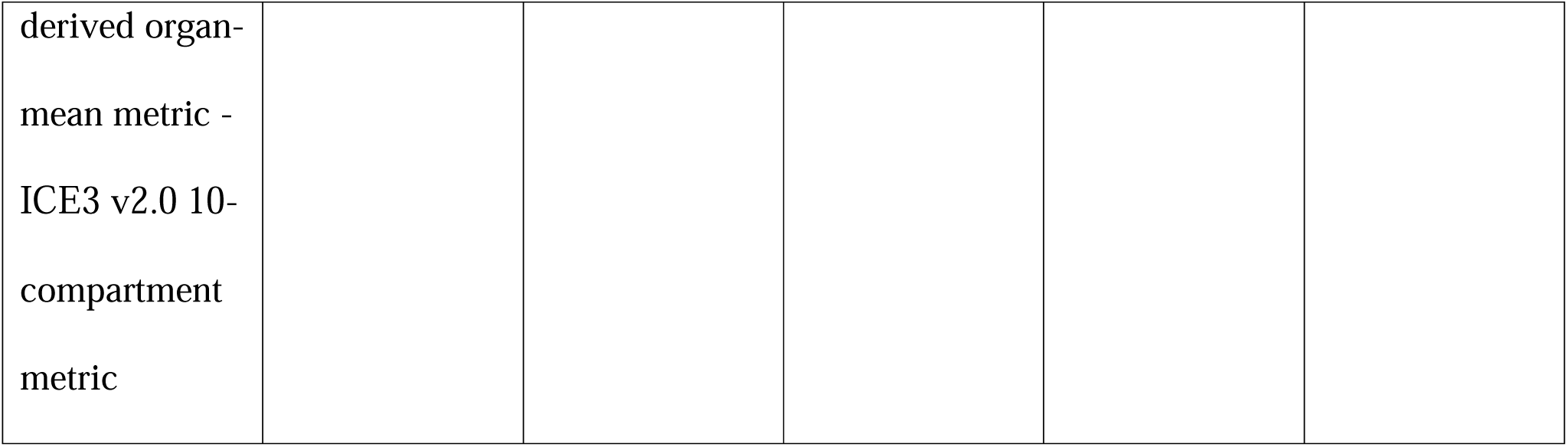
Paired survival bootstrap, 2000 resamples.

In the 88-patient sensitivity set that additionally required baseline ALC and chemotherapy, no exposure reached P<.05; mean lung dose had HR 0.69 (95% CI, 0.48-1.00; P=.052). In the ungated common set, mean heart dose had HR 1.41 (95% CI, 1.04-1.90; P=.026), but the corresponding q value was .158.

### Selected exploratory comparisons

We compared the survival associations of three pairs of dose metrics. These comparisons were chosen after the main analysis and should therefore be considered exploratory.

Mean heart dose showed a more positive association with mortality than ICE3, and this difference remained statistically significant after correcting for the three reported comparisons (difference in standardized log hazard ratios, 0.44; 95% bootstrap CI, 0.13–0.94; adjusted q=.024). The difference between mean heart dose and the HEDOS-derived approximation was smaller and did not remain significant after correction (q=.053). There was no evidence of a meaningful difference between the HEDOS-derived approximation and ICE3 (q=.458).

These findings suggest that mean heart dose and ICE3 may capture different associations with survival, but they do not show that mean heart dose predicts survival more accurately or should replace the other metrics.

### Proportional-Hazards Diagnostics

We tested whether the associations between the dose metrics and survival remained reasonably constant over time, as required by the Cox model. In the 92-patient primary analysis, none of the dose metrics showed evidence of a time-varying effect. The same was true in the 88-patient sensitivity analysis. Baseline ALC showed a possible time-varying association in one mean-lung-dose model (P=.043). Overall, the diagnostics did not identify a violation involving the dose metrics, although they do not confirm that every covariate fully satisfied the proportional-hazards assumption.

### Exploratory analyses of blood-dose distribution and timing

No ICE3-derived kinetic or distributional measure provided additional statistically significant information beyond the ICE3 mean-dose metric after correction for multiple testing. This was true for grade 3 or higher lymphopenia, continuous ALC nadir, overall survival, and the exploratory progression-free-survival analysis.

The most consistent nominal signal involved the zero-cycle fraction, defined as the modeled proportion of circulating blood elements with no recorded passage through an irradiated compartment during the specified simulation interval. A higher zero-cycle fraction was associated with better overall survival (HR, 0.64; P=.016; q=.112) and progression-free survival (HR, 0.70; P=.028; q=.127), but neither association remained significant after false-discovery-rate correction. These findings should therefore be considered hypothesis-generating.

In a separate feasibility analysis of 13 patients, HEDOS blood dose-volume histograms were successfully generated. Introducing 10-second gaps between beams produced only minimal changes in mean blood dose and P98 compared with continuous delivery. Because this analysis was small and based on plan-level organ DVHs rather than beam-specific three-dimensional dose-rate data, the findings should be interpreted only as a limited timing-sensitivity assessment and not as evidence that treatment timing is generally unimportant in full dynamic HEDOS modeling.

## Discussion

A central motivation for this work was the development of a patient-specific digital twin of immune exposure during thoracic radiotherapy. Conventional metrics such as mean heart dose, mean lung dose, and EDIC primarily summarize treatment geometry and organ-level dose. They do not directly describe how circulating lymphocytes move through irradiated cardiac substructures, accumulate dose during repeated passages, or experience heterogeneous dose distributions in spatiotemporal manner over time. Full dynamic frameworks such as HEDOS address some of these processes through blood-flow simulation and time-dependent dose accumulation. However, routine radiotherapy planning datasets generally cover only the treated anatomical region rather than the entire body, limiting fully patient-specific implementation of a whole-body dynamic model in retrospective clinical cohorts. We therefore developed a site-specific prototype (ICE3) that uses routinely available regional anatomy, dose, and treatment information as a practical step toward an immune-exposure digital twin.

The present study evaluated whether the scalar outputs of this prototype provided clinically distinct information before more complex dynamic features were considered. Six scalar dose summaries were compared on common analysis sets, while analyses of blood-dose distribution, circulation, and treatment timing were evaluated separately as exploratory analyses. The principal lymphopenia finding was attenuation rather than disappearance. Adjustment for PTV reduced the magnitude and precision of several associations, and none of the six PTV-adjusted exposure metrics remained significant after correction for multiple testing. The HEDOS-derived organ-mean approximation retained the smallest nominal P value. These results therefore do not establish that simpler metrics are superior or that scalar blood-dose estimates contain no information beyond PTV. Rather, they indicate that much of the signal captured by the evaluated scalar metrics overlaps with treated volume and treatment-field geometry.

PTV adjustment addresses whether a dose metric remains associated with the outcome after accounting for treated volume. Because PTV strongly influences the amount of normal tissue and circulating blood exposed to radiation, it is correlated with several organ- and blood-dose measures. Attenuation after PTV adjustment therefore suggests that part of the observed association is shared with treatment size and geometry. However, this analysis does not establish that PTV directly causes lymphopenia or that it mediates the biological effect of blood irradiation. Moreover, correlation between PTV and the dose metrics can reduce the precision of their estimated independent effects. Accordingly, loss of statistical significance after PTV adjustment should not be interpreted as proof that a metric lacks biological relevance, that PTV is causally dominant, or that the evaluated metrics are equivalent.

After correction for multiple testing, none of the individual dose metrics showed a statistically significant association with overall survival. In exploratory post hoc bootstrap comparisons, the standardized mean-heart-dose coefficient was more positive than the ICE3 coefficient, and this was the only one of three reported contrasts that remained below the Benjamini–Hochberg-adjusted significance threshold. This coefficient difference indicates that the two metrics had different estimated associations with survival, but it does not demonstrate that mean heart dose provides better prediction. The study did not compare model fit, cross-validated discrimination, calibration, clinical utility, or performance in an independent validation cohort, and the Cox models included only limited adjustment for clinical prognostic factors. The mean-heart-dose finding should therefore be considered hypothesis-generating rather than evidence that mean heart dose is superior to, or should replace, more comprehensive immune-exposure metrics.

It is important to distinguish the HEDOS-derived scalar we used in the full cohort from full dynamic HEDOS. The full-cohort variable was calculated from organ mean doses and therefore represented only an approximate mean blood-dose summary. It did not model blood movement or time-dependent dose accumulation and could not evaluate whether blood dose-volume distribution, repeated exposure, dose rate, or treatment timing provided additional information beyond mean-dose metrics. Full dynamic HEDOS was explored in only 13 patients (planning CT simulation is generally limited to the thoracic region and does not contain all whole-body organ information needed for HEDOS). Although blood dose-volume histograms were successfully generated, the analysis did not include beam-specific three-dimensional dose-rate reconstruction and was too small to assess associations with clinical outcomes. These findings should therefore be interpreted as a feasibility analysis rather than a direct comparison of full dynamic HEDOS with ICE3, EDIC, mean heart dose, or mean lung dose.

The exploratory ICE3 kinetic analyses did not show convincing evidence that treatment-timing or recirculation measures added information beyond the ICE3 mean-dose metric. The zero-cycle fraction showed nominal associations with overall and progression-free survival, but these findings did not remain significant after correction for multiple testing and should therefore be considered hypothesis-generating. These results suggest that future development of an immune-exposure digital twin may depend less on further refinement of scalar mean-dose formulas and more on validating dynamic features such as repeated blood passage through irradiated regions, the high-dose tail of the blood-dose distribution, and treatment-delivery timing. Definitive evaluation of these features will require larger cohorts with complete, consistently collected treatment-delivery data such as SBRT, proton therapy, or altered fractionation.

The scan-length analysis identified an important dosimetric quality-control issue. In the original implementation, the body-dose term was calculated from the external contour available on the planning CT and was therefore partly influenced by the anatomical extent of the scan. Differences in scan coverage could alter the metric independently of the patient’s true radiation exposure. After revising the body-compartment definition, the association between the metric and CT scan length was substantially reduced (Figure 3). Future blood-dose studies should clearly define the body region included in the calculation, specify any excluded organs and target volumes, describe the assumed blood-volume scaling, report scan coverage, and evaluate whether the metric is sensitive to differences in image acquisition extent.

**Figure 3.**
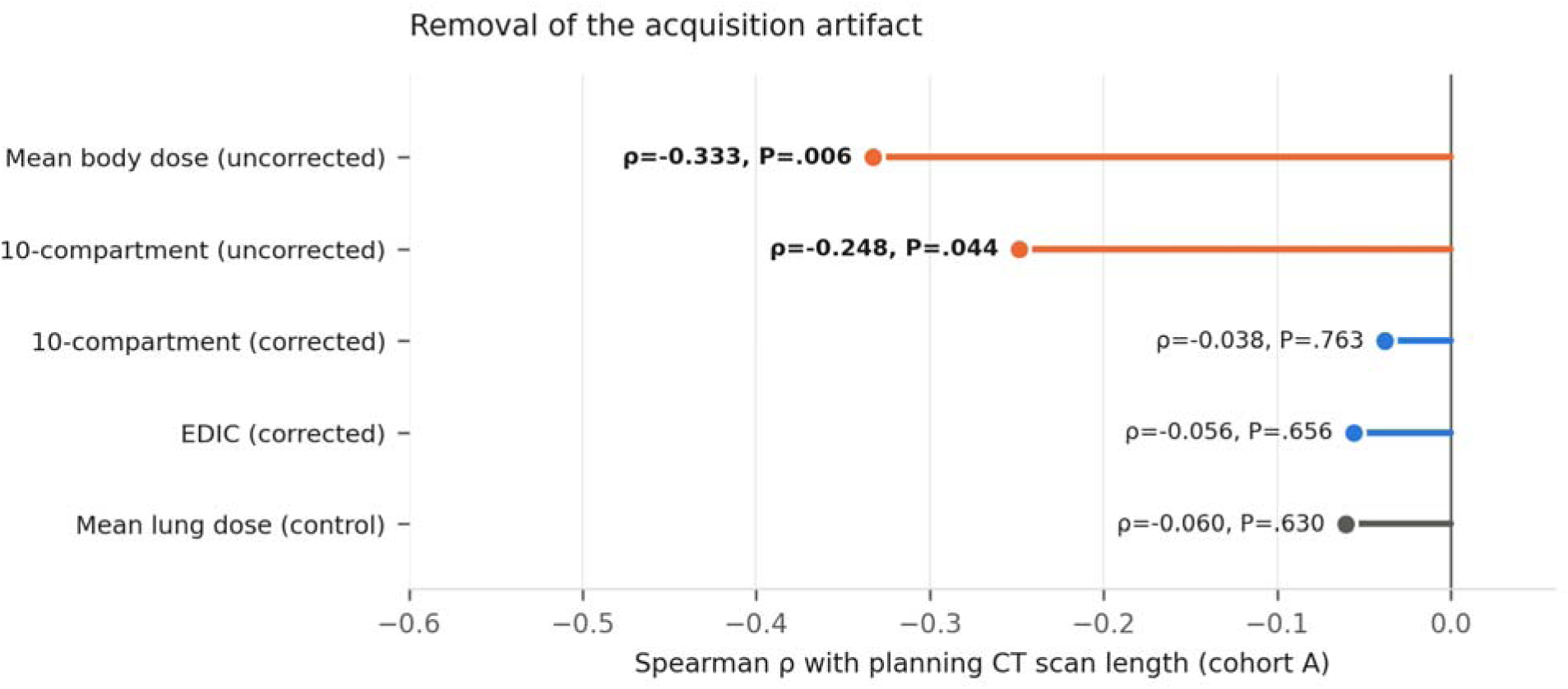
Spearman association between planning CT scan length and body-related dose metrics in Cohort A. Points show observed correlation coefficients with the coefficient and P value labelled; no confidence intervals are displayed. Uncorrected metrics are shown in orange and corrected metrics in blue, with mean lung dose included in grey as a control.

This study has several important limitations. It was retrospective, included two heterogeneous cohorts, and used modest common analysis sets with incomplete clinical adjustment. Several analyses—including the dosimetric gate, selected survival contrasts, progression-free survival, and dynamic modeling—were post hoc, and the lymphopenia model had limited non-events relative to the number of covariates.

The study also evaluated an early, site-specific ICE3 prototype rather than a fully validated immune digital twin. The primary ICE3 endpoint was a scalar mean-dose estimate derived from regional thoracic data under fixed occupancy assumptions; whole-body anatomy, continuous circulation, repeated exposure, and particle-level dose accumulation were not fully modeled. Most treatments were conventionally fractionated thoracic courses of approximately 50–60 Gy in about 25-30 fractions limiting variation in dose per fraction and delivery timing and therefore limiting generalizability to SBRT, hypofractionation, proton therapy, reirradiation, and nonthoracic sites.

Finally, the dynamic HEDOS analysis included only 13 selected cases and lacked beam-specific three-dimensional dose-rate reconstruction, so it could not determine whether full dynamic HEDOS outperforms scalar metrics.

Overall, these findings highlight the limitations of current scalar immune-exposure models without undermining the broader goal of developing a clinically useful immune digital twin. ICE3, EDIC, conventional organ-dose metrics, and the HEDOS-derived mean-dose approximation captured largely overlapping information related to treatment geometry, and none showed a robust association with lymphopenia after adjustment for PTV and correction for multiple testing. A definitive evaluation of the digital-twin concept will require patient-specific dynamic blood-dose modeling in the same cohort, incorporating complete treatment-delivery timing, transparent and reproducible circulation assumptions, other organs such as mediastinal lymph nodes and bone marrow, include lymphocyte replenish model and prospective or external validation.

## Conclusions

In 2 retrospective thoracic radiation cohorts, PTV adjustment attenuated associations between scalar dose summaries and grade 3 or higher lymphopenia, and no PTV-adjusted exposure remained significant after false-discovery-rate correction. No scalar exposure was associated with overall survival on the endpoint-specific common set. One selected post hoc mean-heart-dose-versus-ICE3 coefficient contrast remained below .05 after correction across 3 reported contrasts, but it does not establish superior prediction and requires independent validation. Full dynamic HEDOS was not tested in the primary cohorts; the small bDVH audit remains exploratory.

## Declaration of Generative AI and AI-Assisted Technologies in the Writing Process

During the preparation of this work, the authors used ChatGPT (OpenAI) and Claude to assist with manuscript organization, language editing, consistency auditing, and drafting analysis code. After using this tool, the authors reviewed and edited the content, independently verified all numerical results, and take full responsibility for the content of the publication.

## Competing interests, including any ICE3 intellectual-property or startup interests

University of Kentucky holds intellectual-property of ICE3 software

## Ethics approval, protocol identifiers, and consent waiver

The study was approved by the Medical Ethics Committee of The Fourth Hospital of Hebei Medical University (2026KY165-01).

The study was approved by the Medical Ethics Committee of University of Kentucky IRB Number: 96727 and waver of authorization

## Clinical trial registration

N/A

## License choice for medRxiv

### Author contribution statement

**WY** contributed to software design and engineering, data analysis, and statistical analysis. **YW** performed data extraction and management. **XL** contributed to software application, data extraction, and data processing. **AH** and **JC** contributed to software design and implementation. **DY** contributed to statistics and audit. **MG**, **SZ**, **YZ**, **SY**, **RM**, and **EJ** contributed to data collection. **TO** contributed to data analysis. **EJ** also contributed to data processing. **DC** supervised the study and contributed to manuscript development. **JW** supervised the study and critically reviewed the manuscript. All authors reviewed and approved the final manuscript.

Circulating-blood dose is associated with severe lymphopenia in thoracic radiotherapy, but added simulation complexity provides no detectable incremental information.

## Data Availability

All data produced in the present study are available upon reasonable request to the authors

