## Supplementary material for "Circulating-blood dose is associated with severe lymphopenia in thoracic radiotherapy, but added simulation complexity provides no detectable incremental information": Supplemental_Figure_Legends.md

# Supplemental figure legends

Supplemental Figure S1. Best exported ICE3 v2.0 dynamic or distributional metric by endpoint and test family. Bars show -log10(Benjamini-Hochberg q). The dashed vertical line marks q=.05. No displayed dynamic endpoint family met q<.05.

Supplemental Figure S2. Exploratory HEDOS blood dose-volume histograms (bDVHs) in the 13-patient subset. Thin lines show individual plans under continuous and 10-second inter-beam-gap scenarios; thick lines show scenario medians. This analysis used plan-level organ DVHs and should not be interpreted as full beam-specific 3-dimensional HEDOS.

Supplemental Figure S3. Difference between 10-second inter-beam-gap and continuous HEDOS scenarios for course mean, P98, and maximum blood dose in the 13-patient subset. Points are paired patient-level differences; black markers summarize the mean and 95% confidence interval.

Supplemental Figure S4. Spearman correlations between exported ICE3 v2.0 dynamic metrics and reference quantities. These correlations are descriptive and do not test clinical outcome prediction.
