## Supplementary material for "Circulating-blood dose is associated with severe lymphopenia in thoracic radiotherapy, but added simulation complexity provides no detectable incremental information": V24_Supplementary_Materials.docx

#### S1. v22 Primary Analysis Addendum

### v22 analysis addendum

#### Corrected survival analysis sets

Common survival set requiring all 6 scalar exposures, PTV, cohort, and survival data: n=92, deaths=42.

Sensitivity set additionally requiring baseline ALC and chemotherapy: n=88, deaths=40.

Ungated common survival set: n=94, deaths=44.

#### Disease-site composition in gated main analysis set

| **characteristic** | **Overall** | **Cohort A** | **Cohort B** |
| --- | --- | --- | --- |
| Patients, n | 116 | 66 | 50 |
| Age, median (IQR), y | 69.0 (61.0-76.0) | 75.0 (66.0-77.0) | 64.0 (58.0-69.0) |
| Female sex, n (%) [provisional coding] | 34 (29.3%) | 12 (18.2%) | 22 (44.0%) |
| PTV volume, median (IQR), cc | 286.1 (157.3-410.6) | 258.1 (179.9-364.8) | 303.0 (135.0-445.1) |
| Baseline ALC, median (IQR) | 1.6 (1.1-2.0) | 1.4 (1.1-1.9) | 1.7 (1.3-2.4) |
| Concurrent chemotherapy, n (%) | 71 (61.2%) | 34 (51.5%) | 37 (74.0%) |
| Lung primary, n (%) | 80 (69.0%) | 33 (50.0%) | 47 (94.0%) |
| Esophageal primary, n (%) | 33 (28.4%) | 33 (50.0%) | 0 (0.0%) |
| Other/unknown primary, n (%) | 3 (2.6%) | 0 (0.0%) | 3 (6.0%) |
| Small-cell lung cancer, n (%) | 22 (19.0%) | 0 (0.0%) | 22 (44.0%) |

#### PTV-adjusted survival, common set

| **metric** | **model** | **n** | **events** | **HR** | **lo** | **hi** | **p** | **BH_q** |
| --- | --- | --- | --- | --- | --- | --- | --- | --- |
| ICE3 v2.0 10-compartment circulating-blood metric | PTV-adjusted common survival set | 92 | 42 | 0.848 | 0.579 | 1.241 | 0.395 | 0.592 |
| Corrected EDIC | PTV-adjusted common survival set | 92 | 42 | 1.095 | 0.763 | 1.570 | 0.623 | 0.623 |
| Body-remainder dose | PTV-adjusted common survival set | 92 | 42 | 0.796 | 0.509 | 1.246 | 0.318 | 0.592 |
| HEDOS-derived organ-mean metric | PTV-adjusted common survival set | 92 | 42 | 0.903 | 0.630 | 1.296 | 0.581 | 0.623 |
| Mean lung dose | PTV-adjusted common survival set | 92 | 42 | 0.823 | 0.605 | 1.120 | 0.215 | 0.592 |
| Mean heart dose | PTV-adjusted common survival set | 92 | 42 | 1.312 | 0.960 | 1.795 | 0.089 | 0.531 |

#### Paired survival bootstrap, common set

| **comparison** | **n** | **events** | **observed_dlogHR** | **lo** | **hi** | **p** | **draws** |
| --- | --- | --- | --- | --- | --- | --- | --- |
| Mean heart dose - ICE3 v2.0 10-compartment metric | 92 | 42 | 0.437 | 0.127 | 0.939 | 0.008 | 2000 |
| Mean heart dose - HEDOS-derived organ-mean metric | 92 | 42 | 0.373 | 0.022 | 0.818 | 0.035 | 2000 |
| HEDOS-derived organ-mean metric - ICE3 v2.0 10-compartment metric | 92 | 42 | 0.064 | -0.152 | 0.349 | 0.458 | 2000 |

#### PH diagnostics summary

Main n=92 set: no exposure PH violations; exposure p-values range 0.103-0.877. Minimum model p-value is 0.096.

Sensitivity n=88 set: no exposure PH violations; exposure p-values range 0.313-0.721. Minimum model p-value is 0.043.

The lowest sensitivity-model p-value is for baseline ALC in the Mean lung dose model: p=0.043.

#### S2. PH Diagnostics: Main Survival Set

| **metric** | **model** | **variable** | **n** | **events** | **test_statistic** | **p** | **minus_log2_p** |
| --- | --- | --- | --- | --- | --- | --- | --- |
| ICE3 v2.0 10-compartment circulating-blood metric | PTV-adjusted common survival set | exposure | 92 | 42 | 0.145 | 0.704 | 0.507 |
| ICE3 v2.0 10-compartment circulating-blood metric | PTV-adjusted common survival set | ptv_cc | 92 | 42 | 1.595 | 0.207 | 2.275 |
| Corrected EDIC | PTV-adjusted common survival set | exposure | 92 | 42 | 0.049 | 0.826 | 0.276 |
| Corrected EDIC | PTV-adjusted common survival set | ptv_cc | 92 | 42 | 1.837 | 0.175 | 2.512 |
| Body-remainder dose | PTV-adjusted common survival set | exposure | 92 | 42 | 0.113 | 0.737 | 0.441 |
| Body-remainder dose | PTV-adjusted common survival set | ptv_cc | 92 | 42 | 2.214 | 0.137 | 2.871 |
| HEDOS-derived organ-mean metric | PTV-adjusted common survival set | exposure | 92 | 42 | 0.090 | 0.764 | 0.389 |
| HEDOS-derived organ-mean metric | PTV-adjusted common survival set | ptv_cc | 92 | 42 | 0.978 | 0.323 | 1.631 |
| Mean lung dose | PTV-adjusted common survival set | exposure | 92 | 42 | 2.663 | 0.103 | 3.283 |
| Mean lung dose | PTV-adjusted common survival set | ptv_cc | 92 | 42 | 2.765 | 0.096 | 3.375 |
| Mean heart dose | PTV-adjusted common survival set | exposure | 92 | 42 | 0.024 | 0.877 | 0.189 |
| Mean heart dose | PTV-adjusted common survival set | ptv_cc | 92 | 42 | 2.075 | 0.150 | 2.740 |

#### S3. PH Diagnostics: Sensitivity Survival Set

| **metric** | **model** | **variable** | **n** | **events** | **test_statistic** | **p** | **minus_log2_p** |
| --- | --- | --- | --- | --- | --- | --- | --- |
| ICE3 v2.0 10-compartment circulating-blood metric | PTV + baseline ALC + chemotherapy | alc_baseline | 88 | 40 | 3.134 | 0.077 | 3.705 |
| ICE3 v2.0 10-compartment circulating-blood metric | PTV + baseline ALC + chemotherapy | conc_chemo | 88 | 40 | 0.372 | 0.542 | 0.884 |
| ICE3 v2.0 10-compartment circulating-blood metric | PTV + baseline ALC + chemotherapy | exposure | 88 | 40 | 1.018 | 0.313 | 1.676 |
| ICE3 v2.0 10-compartment circulating-blood metric | PTV + baseline ALC + chemotherapy | ptv_cc | 88 | 40 | 0.059 | 0.808 | 0.307 |
| Corrected EDIC | PTV + baseline ALC + chemotherapy | alc_baseline | 88 | 40 | 2.396 | 0.122 | 3.039 |
| Corrected EDIC | PTV + baseline ALC + chemotherapy | conc_chemo | 88 | 40 | 1.007 | 0.316 | 1.664 |
| Corrected EDIC | PTV + baseline ALC + chemotherapy | exposure | 88 | 40 | 0.441 | 0.506 | 0.982 |
| Corrected EDIC | PTV + baseline ALC + chemotherapy | ptv_cc | 88 | 40 | 0.101 | 0.750 | 0.414 |
| Body-remainder dose | PTV + baseline ALC + chemotherapy | alc_baseline | 88 | 40 | 2.831 | 0.092 | 3.435 |
| Body-remainder dose | PTV + baseline ALC + chemotherapy | conc_chemo | 88 | 40 | 0.645 | 0.422 | 1.245 |
| Body-remainder dose | PTV + baseline ALC + chemotherapy | exposure | 88 | 40 | 0.698 | 0.403 | 1.310 |
| Body-remainder dose | PTV + baseline ALC + chemotherapy | ptv_cc | 88 | 40 | 0.002 | 0.964 | 0.054 |
| HEDOS-derived organ-mean metric | PTV + baseline ALC + chemotherapy | alc_baseline | 88 | 40 | 1.904 | 0.168 | 2.576 |
| HEDOS-derived organ-mean metric | PTV + baseline ALC + chemotherapy | conc_chemo | 88 | 40 | 0.506 | 0.477 | 1.068 |
| HEDOS-derived organ-mean metric | PTV + baseline ALC + chemotherapy | exposure | 88 | 40 | 0.128 | 0.721 | 0.473 |
| HEDOS-derived organ-mean metric | PTV + baseline ALC + chemotherapy | ptv_cc | 88 | 40 | 0.030 | 0.863 | 0.212 |
| Mean lung dose | PTV + baseline ALC + chemotherapy | alc_baseline | 88 | 40 | 4.101 | 0.043 | 4.544 |
| Mean lung dose | PTV + baseline ALC + chemotherapy | conc_chemo | 88 | 40 | 1.057 | 0.304 | 1.718 |
| Mean lung dose | PTV + baseline ALC + chemotherapy | exposure | 88 | 40 | 0.793 | 0.373 | 1.422 |
| Mean lung dose | PTV + baseline ALC + chemotherapy | ptv_cc | 88 | 40 | 0.139 | 0.710 | 0.495 |
| Mean heart dose | PTV + baseline ALC + chemotherapy | alc_baseline | 88 | 40 | 1.563 | 0.211 | 2.243 |
| Mean heart dose | PTV + baseline ALC + chemotherapy | conc_chemo | 88 | 40 | 0.735 | 0.391 | 1.353 |
| Mean heart dose | PTV + baseline ALC + chemotherapy | exposure | 88 | 40 | 0.741 | 0.389 | 1.361 |
| Mean heart dose | PTV + baseline ALC + chemotherapy | ptv_cc | 88 | 40 | 0.436 | 0.509 | 0.974 |

#### S4. Exported ICE3 v2.0 Kinetic and Distributional Audit

### ICE3 v2.0 Dynamic-Metric Audit

#### Scope

This rerun uses `results_v19/analysis_frame_v19.csv`, which is the exported dose/contour-derived analysis frame in the thoracic package. The external-review zip does not contain raw DICOM dose grids or RTSTRUCT contours, so this is not a fresh autosegmentation or full HEDOS particle-flow run.

The audit tests the ICE3 v2.0 dynamic/kinetic columns already present in the frame and asks whether they add clinical signal beyond the corrected ICE3 v2.0 mean-dose metric used in the locked scalar analysis.

Main-analysis records: 121. Passing locked dose/mask gate: 116.

#### Available Dynamic ICE3 v2.0 Columns

| **column** | **metric** | **analysis-gated n** | **median** | **IQR** |
| --- | --- | --- | --- | --- |
| `dmax_wc` | ICE3 v2.0 Dmax, whole course | 116 | 14.611 | 10.479-18.125 |
| `d100_wc` | ICE3 v2.0 D100/minimum particle dose, whole course | 116 | 5.099 | 3.575-6.501 |
| `d100_perfx_Gy` | ICE3 v2.0 D100/minimum particle dose, per fraction | 116 | 0.172 | 0.125-0.221 |
| `dose_per_hit` | Mean dose per exposure event | 116 | 0.160 | 0.095-0.200 |
| `exposure_cycles` | Mean exposure cycles | 116 | 1.248 | 0.996-1.745 |
| `zero_cycle_frac` | Zero-cycle fraction | 116 | 0.238 | 0.135-0.319 |
| `beam_on_s` | Beam-on time | 103 | 76.500 | 61.930-119.330 |

#### Internal Structure Checks

| **check** | **n** | **Pearson** | **Spearman** | **max absolute error** |
| --- | --- | --- | --- | --- |
| zero_cycle_frac vs exp(-exposure_cycles) | 116 | 0.99849 | -1.00000 | 0.05114 |
| dose_per_hit * exposure_cycles vs ICE3 v2.0 mean per fraction | 116 | 0.88896 | 0.88714 | 0.1889 |
| exposure_cycles vs beam_on_s | 103 | 0.81244 | 0.87408 | - |

#### Correlation With Corrected ICE3 v2.0 Mean

| **metric** | **n** | **Spearman rho** |
| --- | --- | --- |
| ICE3 v2.0 Dmax, whole course | 116 | 0.763 |
| ICE3 v2.0 D100/minimum particle dose, whole course | 116 | 0.863 |
| ICE3 v2.0 D100/minimum particle dose, per fraction | 116 | 0.844 |
| Mean dose per exposure event | 116 | 0.769 |
| Mean exposure cycles | 116 | 0.083 |
| Zero-cycle fraction | 116 | -0.083 |
| Beam-on time | 103 | -0.028 |

#### Outcome Models, Maximal Available Analysis-Gated Set

Grade ≥3 lymphopenia models adjust for baseline ALC, concurrent chemotherapy, and cohort. Cox models are stratified by cohort. The PTV column adds ptv_cc. IMPORTANT: the rows of this table are NOT all fitted on the same patients. This audit used the maximal set available for each individual metric, so n ranges from 94 to 105 patients (lymphopenia events 69 to 76) and from 92 to 99 patients (42 to 51 deaths) for overall survival. Estimates in this table are therefore not directly comparable across rows and differ from the main-text Tables 2 and 3A, which are restricted to the common analysis sets (n=94 and n=92) in which every exposure is available on identical patients. Per-row sample sizes are given in dynamic_metric_outcome_models.csv.

| **metric** | **G3+ base OR/SD** | **G3+ +PTV OR/SD** | **OS base HR/SD** | **OS +PTV HR/SD** |
| --- | --- | --- | --- | --- |
| ICE3 v2.0 corrected mean-dose metric | 2.86 (1.48-5.53; P=0.002) | 2.29 (1.05-4.99; P=0.036) | 1.23 (0.86-1.75; P=0.253) | 0.92 (0.63-1.34; P=0.667) |
| HEDOS-derived mean-dose metric | 3.47 (1.70-7.08; P=<0.001) | 2.54 (1.18-5.45; P=0.017) | 1.13 (0.80-1.60; P=0.493) | 0.90 (0.63-1.30; P=0.581) |
| Mean heart dose | 1.73 (1.00-2.99; P=0.050) | 1.77 (0.92-3.39; P=0.086) | 1.44 (1.09-1.90; P=0.011) | 1.39 (1.04-1.85; P=0.028) |
| Mean lung dose | 2.31 (1.27-4.19; P=0.006) | 2.11 (1.00-4.43; P=0.049) | 1.12 (0.86-1.47; P=0.397) | 0.90 (0.66-1.21; P=0.478) |
| ICE3 v2.0 Dmax, whole course | 1.39 (0.86-2.25; P=0.184) | 1.32 (0.77-2.29; P=0.314) | 0.88 (0.67-1.16; P=0.384) | 0.78 (0.57-1.08; P=0.131) |
| ICE3 v2.0 D100/minimum particle dose, whole course | 2.25 (1.17-4.33; P=0.015) | 1.60 (0.73-3.52; P=0.238) | 1.34 (0.92-1.94; P=0.124) | 1.08 (0.73-1.62; P=0.690) |
| ICE3 v2.0 D100/minimum particle dose, per fraction | 2.54 (1.31-4.92; P=0.006) | 1.89 (0.89-4.03; P=0.100) | 1.27 (0.89-1.81; P=0.185) | 1.06 (0.72-1.56; P=0.779) |
| Mean dose per exposure event | 1.91 (1.04-3.52; P=0.038) | 1.63 (0.80-3.33; P=0.179) | 0.92 (0.65-1.31; P=0.656) | 0.76 (0.50-1.16; P=0.209) |
| Mean exposure cycles | 1.14 (0.70-1.85; P=0.589) | 1.03 (0.57-1.87; P=0.920) | 1.32 (1.04-1.68; P=0.025) | 1.33 (0.99-1.79; P=0.057) |
| Zero-cycle fraction | 0.69 (0.43-1.10; P=0.123) | 0.81 (0.47-1.42; P=0.471) | 0.66 (0.48-0.89; P=0.006) | 0.68 (0.48-0.96; P=0.030) |
| Beam-on time | 1.36 (0.74-2.51; P=0.317) | 1.14 (0.58-2.24; P=0.699) | 1.08 (0.82-1.44; P=0.580) | 1.09 (0.80-1.47; P=0.590) |

#### Incremental Tests Beyond Corrected ICE3 v2.0 Mean

Each row tests the residual part of the dynamic metric after regressing it on `ice3_v18`, with `ice3_v18` retained in the outcome model.

| **metric residual** | **G3+ base P** | **G3+ +PTV P** | **OS base P** | **OS +PTV P** |
| --- | --- | --- | --- | --- |
| ICE3 v2.0 Dmax, whole course | 0.161 | 0.593 | 0.031 | 0.094 |
| ICE3 v2.0 D100/minimum particle dose, whole course | 0.715 | 0.616 | 0.314 | 0.326 |
| ICE3 v2.0 D100/minimum particle dose, per fraction | 0.673 | 0.784 | 0.465 | 0.411 |
| Mean dose per exposure event | 0.684 | 0.977 | 0.163 | 0.215 |
| Mean exposure cycles | 0.781 | 0.916 | 0.024 | 0.049 |
| Zero-cycle fraction | 0.388 | 0.766 | 0.009 | 0.016 |
| Beam-on time | 0.626 | 0.926 | 0.607 | 0.394 |

#### OS Sensitivity for Nominal Timing Signals

These Cox sensitivity models are stratified by cohort. The clinical model adds PTV, age, stage, and concurrent chemotherapy. The beam-on model adds PTV and beam-on time to test whether timing summaries are just delivery-time surrogates.

| **metric** | **primary +clinical** | **residual +clinical** | **primary +beam-on** | **residual +beam-on** |
| --- | --- | --- | --- | --- |
| ICE3 v2.0 Dmax, whole course | 0.79 (0.54-1.15; P=0.220, q=0.514) | 0.65 (0.37-1.16; P=0.147, q=0.432) | 0.73 (0.53-1.02; P=0.063, q=0.148) | 0.68 (0.41-1.11; P=0.124, q=0.248) |
| ICE3 v2.0 D100/minimum particle dose, whole course | 1.08 (0.69-1.70; P=0.735, q=0.735) | 1.29 (0.67-2.49; P=0.451, q=0.472) | 1.03 (0.68-1.57; P=0.881, q=0.984) | 1.45 (0.82-2.56; P=0.207, q=0.248) |
| ICE3 v2.0 D100/minimum particle dose, per fraction | 1.08 (0.70-1.68; P=0.727, q=0.735) | 1.30 (0.68-2.47; P=0.429, q=0.472) | 1.00 (0.66-1.52; P=0.984, q=0.984) | 1.36 (0.78-2.39; P=0.282, q=0.282) |
| Mean dose per exposure event | 0.82 (0.52-1.29; P=0.383, q=0.669) | 0.77 (0.44-1.35; P=0.366, q=0.472) | 0.68 (0.43-1.07; P=0.092, q=0.148) | 0.69 (0.40-1.18; P=0.174, q=0.248) |
| Mean exposure cycles | 1.24 (0.89-1.72; P=0.198, q=0.514) | 1.25 (0.90-1.72; P=0.185, q=0.432) | 1.80 (0.90-3.62; P=0.099, q=0.148) | 1.87 (0.86-4.05; P=0.113, q=0.248) |
| Zero-cycle fraction | 0.75 (0.52-1.07; P=0.109, q=0.514) | 0.72 (0.50-1.04; P=0.083, q=0.432) | 0.57 (0.34-0.95; P=0.031, q=0.148) | 0.45 (0.22-0.90; P=0.023, q=0.141) |
| Beam-on time | 1.07 (0.73-1.58; P=0.723, q=0.735) | 1.17 (0.77-1.77; P=0.472, q=0.472) | - | - |

#### Common Dynamic Complete-Case Sensitivity

Complete case for corrected ICE3 v2.0 mean plus all dynamic columns, PTV, and OS fields: n=84, G3+ events=62, deaths=38.

| **metric** | **G3+ +PTV** | **OS +PTV** |
| --- | --- | --- |
| ICE3 v2.0 corrected mean-dose metric | 2.16 (0.98-4.75; P=0.055) | 0.74 (0.49-1.13; P=0.170) |
| ICE3 v2.0 Dmax, whole course | 1.56 (0.81-2.98; P=0.182) | 0.68 (0.48-0.97; P=0.035) |
| ICE3 v2.0 D100/minimum particle dose, whole course | 1.70 (0.75-3.82; P=0.202) | 1.02 (0.66-1.58; P=0.938) |
| ICE3 v2.0 D100/minimum particle dose, per fraction | 1.97 (0.91-4.30; P=0.087) | 0.97 (0.63-1.49; P=0.898) |
| Mean dose per exposure event | 1.94 (0.86-4.34; P=0.109) | 0.67 (0.43-1.04; P=0.075) |
| Mean exposure cycles | 0.89 (0.45-1.75; P=0.734) | 1.49 (1.05-2.12; P=0.027) |
| Zero-cycle fraction | 0.86 (0.48-1.55; P=0.611) | 0.62 (0.41-0.94; P=0.025) |
| Beam-on time | 0.99 (0.49-1.98; P=0.968) | 1.09 (0.80-1.50; P=0.587) |

#### Interpretation

No ICE3 v2.0 dynamic/kinetic summary in the locked exported analysis frame showed FDR-significant incremental outcome information beyond corrected ICE3 v2.0 mean dose after PTV adjustment.

There is a nominal OS timing/distribution signal after PTV adjustment (Mean exposure cycles residual HR 1.34, P=0.049, q=0.173; Zero-cycle fraction residual HR 0.64, P=0.016, q=0.112), but it does not survive FDR correction and attenuates in the clinical sensitivity models.

This scalar-export audit alone does not prove that full dynamic HEDOS-style particle dose distributions are uninformative. The separate HEDOS-DVH subset audit evaluates locally available DVH/RTPLAN cases, but the full locked analysis frame still lacks particle-level dose histories.

This comparison should therefore be read as a test of scalar mean-dose summaries and exported ICE3 v2.0 kinetic summaries, and not as evidence that dynamic blood-flow dose modelling has no value.

#### S5. v23 ICE3 v2.0 Dynamic Endpoint Audit

### v23 ICE3 v2.0 Dynamic Endpoint Audit

#### Scope

This addendum tests exported ICE3 v2.0 kinetic and distributional summaries already present in `results_v19/analysis_frame_v19.csv`. It does not rerun ICE3 v2.0 or HEDOS and should be treated as exploratory.

Analysis-gated records: 116.

#### Endpoint-Level Results

##### G3+ lymphopenia

**primary**

Sample/event range: n=94-100; events=69-73.

| **metric** | **kind** | **estimate** | **lo** | **hi** | **p** | **q** |
| --- | --- | --- | --- | --- | --- | --- |
| ICE3 v2.0 D100/minimum particle dose, per fraction | OR | 1.889 | 0.885 | 4.033 | 0.100 | 0.550 |
| Mean dose per exposure event | OR | 1.632 | 0.798 | 3.334 | 0.179 | 0.550 |
| ICE3 v2.0 D100/minimum particle dose, whole course | OR | 1.605 | 0.731 | 3.522 | 0.238 | 0.550 |
| ICE3 v2.0 Dmax, whole course | OR | 1.324 | 0.767 | 2.286 | 0.314 | 0.550 |
| Zero-cycle fraction | OR | 0.815 | 0.467 | 1.422 | 0.471 | 0.660 |
| Beam-on time | OR | 1.142 | 0.583 | 2.237 | 0.699 | 0.816 |
| Mean exposure cycles | OR | 1.031 | 0.569 | 1.870 | 0.920 | 0.920 |

**residual beyond ICE3 v2.0 mean**

Sample/event range: n=94-100; events=69-73.

| **metric** | **kind** | **estimate** | **lo** | **hi** | **p** | **q** |
| --- | --- | --- | --- | --- | --- | --- |
| ICE3 v2.0 Dmax, whole course | OR | 0.823 | 0.404 | 1.679 | 0.593 | 0.977 |
| ICE3 v2.0 D100/minimum particle dose, whole course | OR | 0.747 | 0.239 | 2.332 | 0.616 | 0.977 |
| Zero-cycle fraction | OR | 0.917 | 0.517 | 1.625 | 0.766 | 0.977 |
| ICE3 v2.0 D100/minimum particle dose, per fraction | OR | 1.156 | 0.409 | 3.264 | 0.784 | 0.977 |
| Mean exposure cycles | OR | 0.967 | 0.516 | 1.813 | 0.916 | 0.977 |
| Beam-on time | OR | 1.034 | 0.512 | 2.088 | 0.926 | 0.977 |
| Mean dose per exposure event | OR | 1.013 | 0.406 | 2.531 | 0.977 | 0.977 |

##### ALC nadir

**primary**

Sample size range: n=94-100.

| **metric** | **kind** | **estimate** | **lo** | **hi** | **p** | **q** |
| --- | --- | --- | --- | --- | --- | --- |
| ICE3 v2.0 D100/minimum particle dose, per fraction | beta | -0.052 | -0.101 | -0.003 | 0.038 | 0.111 |
| ICE3 v2.0 Dmax, whole course | beta | -0.043 | -0.085 | -0.001 | 0.043 | 0.111 |
| Mean dose per exposure event | beta | -0.051 | -0.103 | 0.002 | 0.058 | 0.111 |
| ICE3 v2.0 D100/minimum particle dose, whole course | beta | -0.047 | -0.097 | 0.003 | 0.064 | 0.111 |
| Beam-on time | beta | -0.025 | -0.081 | 0.031 | 0.383 | 0.536 |
| Zero-cycle fraction | beta | 0.014 | -0.032 | 0.059 | 0.547 | 0.638 |
| Mean exposure cycles | beta | -0.004 | -0.052 | 0.045 | 0.883 | 0.883 |

**residual beyond ICE3 v2.0 mean**

Sample size range: n=94-100.

| **metric** | **kind** | **estimate** | **lo** | **hi** | **p** | **q** |
| --- | --- | --- | --- | --- | --- | --- |
| ICE3 v2.0 D100/minimum particle dose, whole course | beta | 0.025 | -0.049 | 0.099 | 0.502 | 0.956 |
| Beam-on time | beta | -0.013 | -0.067 | 0.042 | 0.642 | 0.956 |
| ICE3 v2.0 D100/minimum particle dose, per fraction | beta | 0.009 | -0.062 | 0.079 | 0.809 | 0.956 |
| ICE3 v2.0 Dmax, whole course | beta | 0.005 | -0.052 | 0.062 | 0.860 | 0.956 |
| Mean dose per exposure event | beta | 0.005 | -0.063 | 0.073 | 0.878 | 0.956 |
| Zero-cycle fraction | beta | -0.002 | -0.046 | 0.043 | 0.937 | 0.956 |
| Mean exposure cycles | beta | 0.001 | -0.045 | 0.048 | 0.956 | 0.956 |

##### Overall survival

**primary**

Sample/event range: n=92-99; events=42-47.

| **metric** | **kind** | **estimate** | **lo** | **hi** | **p** | **q** |
| --- | --- | --- | --- | --- | --- | --- |
| Zero-cycle fraction | HR | 0.680 | 0.481 | 0.962 | 0.030 | 0.199 |
| Mean exposure cycles | HR | 1.332 | 0.992 | 1.790 | 0.057 | 0.199 |
| ICE3 v2.0 Dmax, whole course | HR | 0.780 | 0.566 | 1.077 | 0.131 | 0.306 |
| Mean dose per exposure event | HR | 0.763 | 0.500 | 1.164 | 0.209 | 0.366 |
| Beam-on time | HR | 1.087 | 0.803 | 1.472 | 0.590 | 0.779 |
| ICE3 v2.0 D100/minimum particle dose, whole course | HR | 1.085 | 0.727 | 1.619 | 0.690 | 0.779 |
| ICE3 v2.0 D100/minimum particle dose, per fraction | HR | 1.057 | 0.717 | 1.559 | 0.779 | 0.779 |

**residual beyond ICE3 v2.0 mean**

Sample/event range: n=92-99; events=42-47.

| **metric** | **kind** | **estimate** | **lo** | **hi** | **p** | **q** |
| --- | --- | --- | --- | --- | --- | --- |
| Zero-cycle fraction | HR | 0.644 | 0.451 | 0.921 | 0.016 | 0.112 |
| Mean exposure cycles | HR | 1.339 | 1.001 | 1.792 | 0.049 | 0.173 |
| ICE3 v2.0 Dmax, whole course | HR | 0.647 | 0.388 | 1.077 | 0.094 | 0.219 |
| Mean dose per exposure event | HR | 0.731 | 0.445 | 1.200 | 0.215 | 0.376 |
| ICE3 v2.0 D100/minimum particle dose, whole course | HR | 1.323 | 0.757 | 2.313 | 0.326 | 0.411 |
| Beam-on time | HR | 1.151 | 0.833 | 1.591 | 0.394 | 0.411 |
| ICE3 v2.0 D100/minimum particle dose, per fraction | HR | 1.253 | 0.732 | 2.145 | 0.411 | 0.411 |

##### Progression-free survival

**primary**

Sample/event range: n=94-100; events=54-58.

| **metric** | **kind** | **estimate** | **lo** | **hi** | **p** | **q** |
| --- | --- | --- | --- | --- | --- | --- |
| Mean exposure cycles | HR | 1.342 | 1.009 | 1.785 | 0.043 | 0.162 |
| Zero-cycle fraction | HR | 0.736 | 0.545 | 0.995 | 0.046 | 0.162 |
| Mean dose per exposure event | HR | 0.745 | 0.508 | 1.092 | 0.132 | 0.307 |
| ICE3 v2.0 Dmax, whole course | HR | 0.824 | 0.619 | 1.097 | 0.185 | 0.324 |
| Beam-on time | HR | 1.174 | 0.901 | 1.528 | 0.235 | 0.328 |
| ICE3 v2.0 D100/minimum particle dose, per fraction | HR | 0.960 | 0.676 | 1.365 | 0.821 | 0.866 |
| ICE3 v2.0 D100/minimum particle dose, whole course | HR | 0.969 | 0.675 | 1.393 | 0.866 | 0.866 |

**residual beyond ICE3 v2.0 mean**

Sample/event range: n=94-100; events=54-58.

| **metric** | **kind** | **estimate** | **lo** | **hi** | **p** | **q** |
| --- | --- | --- | --- | --- | --- | --- |
| Zero-cycle fraction | HR | 0.703 | 0.514 | 0.962 | 0.028 | 0.127 |
| Mean exposure cycles | HR | 1.353 | 1.019 | 1.796 | 0.036 | 0.127 |
| Mean dose per exposure event | HR | 0.693 | 0.439 | 1.095 | 0.116 | 0.199 |
| Beam-on time | HR | 1.243 | 0.934 | 1.655 | 0.136 | 0.199 |
| ICE3 v2.0 Dmax, whole course | HR | 0.720 | 0.465 | 1.116 | 0.142 | 0.199 |
| ICE3 v2.0 D100/minimum particle dose, whole course | HR | 1.040 | 0.627 | 1.722 | 0.880 | 0.943 |
| ICE3 v2.0 D100/minimum particle dose, per fraction | HR | 1.018 | 0.624 | 1.659 | 0.943 | 0.943 |

#### Interpretation

No ICE3 v2.0 dynamic or kinetic metric survived false-discovery-rate control for G3+ lymphopenia, continuous ALC nadir, OS, or post hoc PFS. Dose-distribution summaries such as Dmax, D100, and dose per exposure event were not useful as incremental clinical predictors beyond ICE3 v2.0 mean dose. The only repeated pattern was a nominal timing/recirculation signal for exposure cycles and zero-cycle fraction in OS and PFS, but q values remained above .05 and these variables are vulnerable to delivery-time and treatment-complexity confounding. These results are hypothesis-generating and should not be promoted to a primary finding.

#### S6. Exploratory HEDOS-DVH Subset Audit

### v21 HEDOS Dynamic DVH Subset Audit

This audit uses the official HEDOS flow model and DVH-based particle-dose accumulation on locally available full-DVH exports from one participating institution.

#### Scope

- Available v19 AI-labelled cases screened: 13
- Cases with usable DVH + RTPLAN dynamic HEDOS outputs: 13
- HEDOS particles per plan-scenario: 10,000
- Time step: 0.05 s
- Gapped scenario inter-beam gap: 10 s

#### Interpretation Guardrails

- These outputs are dynamic HEDOS blood-particle dose distributions and bDVHs, not scalar mean-only HEDOS-derived summaries.
- They are HEDOS-DVH approximations because plan-level organ DVHs are reused across beams; beam-specific 3D dose-rate fields were not reconstructed.
- The overlap with the v19 outcome frame is too small for primary clinical inference.

#### Files

- `hedos_dynamic_subset_metrics.csv`: case-level course bDVH summary metrics.
- `hedos_dynamic_plan_metrics.csv`: plan-level fraction and course metrics.
- `hedos_dynamic_bdvhs.csv`: bDVH curves for each case and timing scenario.
- `hedos_dynamic_correlations.csv`: descriptive Spearman correlations with v19 scalar metrics and outcomes.
- hedos_dynamic_correlations.csv: descriptive Spearman correlations with v19 scalar metrics and outcomes. (A missing-input audit file, hedos_dynamic_skipped.csv, is referenced by the analysis code but is not distributed with this package; all 13 screened cases yielded usable output, so the file is empty.)

#### Scenario Summary

```text

scenario	n	mean_course_mean_Gy	median_course_p98_Gy	median_V1Gy_pct	median_V5Gy_pct

scenario	n	mean_course_mean_Gy	median_course_p98_Gy	median_V1Gy_pct	median_V5Gy_pct
continuous	13	3.266	3.684	100.0	0.0
gapped	13	3.281	3.673	100.0	0.0

gapped	13	3.280519943559244	3.6726255296955324	100.0	0.0

```

#### Top Descriptive Correlations

```text

scenario	metric	reference	n	spearman_rho	p_value

continuous	hedos_course_mean_Gy	v19_mld_Gy	13	0.8791208791208791	7.544562359359131e-05

gapped	hedos_course_median_Gy	v19_mld_Gy	13	0.8791208791208791	7.544562359359131e-05

gapped	hedos_course_mean_Gy	v19_mld_Gy	13	0.8791208791208791	7.544562359359131e-05

continuous	hedos_course_median_Gy	v19_mld_Gy	13	0.8681318681318682	0.0001191025598004272

continuous	hedos_course_p10_Gy	v19_mld_Gy	13	0.8626373626373626	0.00014744785947619777

gapped	hedos_course_p90_Gy	v19_mld_Gy	13	0.8626373626373626	0.00014744785947619777

gapped	hedos_course_p10_Gy	v19_mld_Gy	13	0.8626373626373626	0.00014744785947619777

continuous	hedos_course_p90_Gy	v19_mld_Gy	13	0.8626373626373626	0.00014744785947619777

gapped	hedos_course_p98_Gy	v19_mld_Gy	13	0.8571428571428571	0.00018093408872524061

gapped	hedos_course_max_Gy	v19_mld_Gy	13	0.8516483516483516	0.00022021941625366217

continuous	hedos_course_p98_Gy	v19_mld_Gy	13	0.8516483516483516	0.00022021941625366217

continuous	hedos_course_max_Gy	v19_mld_Gy	13	0.8461538461538461	0.00026601205575225306

continuous	hedos_course_max_Gy	v19_mhd_Gy	13	0.7857142857142857	0.0014541896038438233

gapped	hedos_course_mean_Gy	v19_ice3_v18_Gy	13	0.7802197802197801	0.0016525006574429557

gapped	hedos_course_median_Gy	v19_ice3_v18_Gy	13	0.7802197802197801	0.0016525006574429557

continuous	hedos_course_mean_Gy	v19_ice3_v18_Gy	13	0.7802197802197801	0.0016525006574429557

gapped	hedos_course_max_Gy	v19_mhd_Gy	13	0.7802197802197801	0.0016525006574429557

continuous	hedos_course_p90_Gy	v19_ice3_v18_Gy	13	0.7747252747252747	0.0018713615028719084

continuous	hedos_course_max_Gy	v19_ice3_v18_Gy	13	0.7747252747252747	0.0018713615028719084

gapped	hedos_course_p90_Gy	v19_ice3_v18_Gy	13	0.7747252747252747	0.0018713615028719084

```
