## Supplementary figures and images for "Circulating-blood dose is associated with severe lymphopenia in thoracic radiotherapy, but added simulation complexity provides no detectable incremental information"

### Supplemental_Figure_S1_dynamic_endpoint_qvalues.jpg

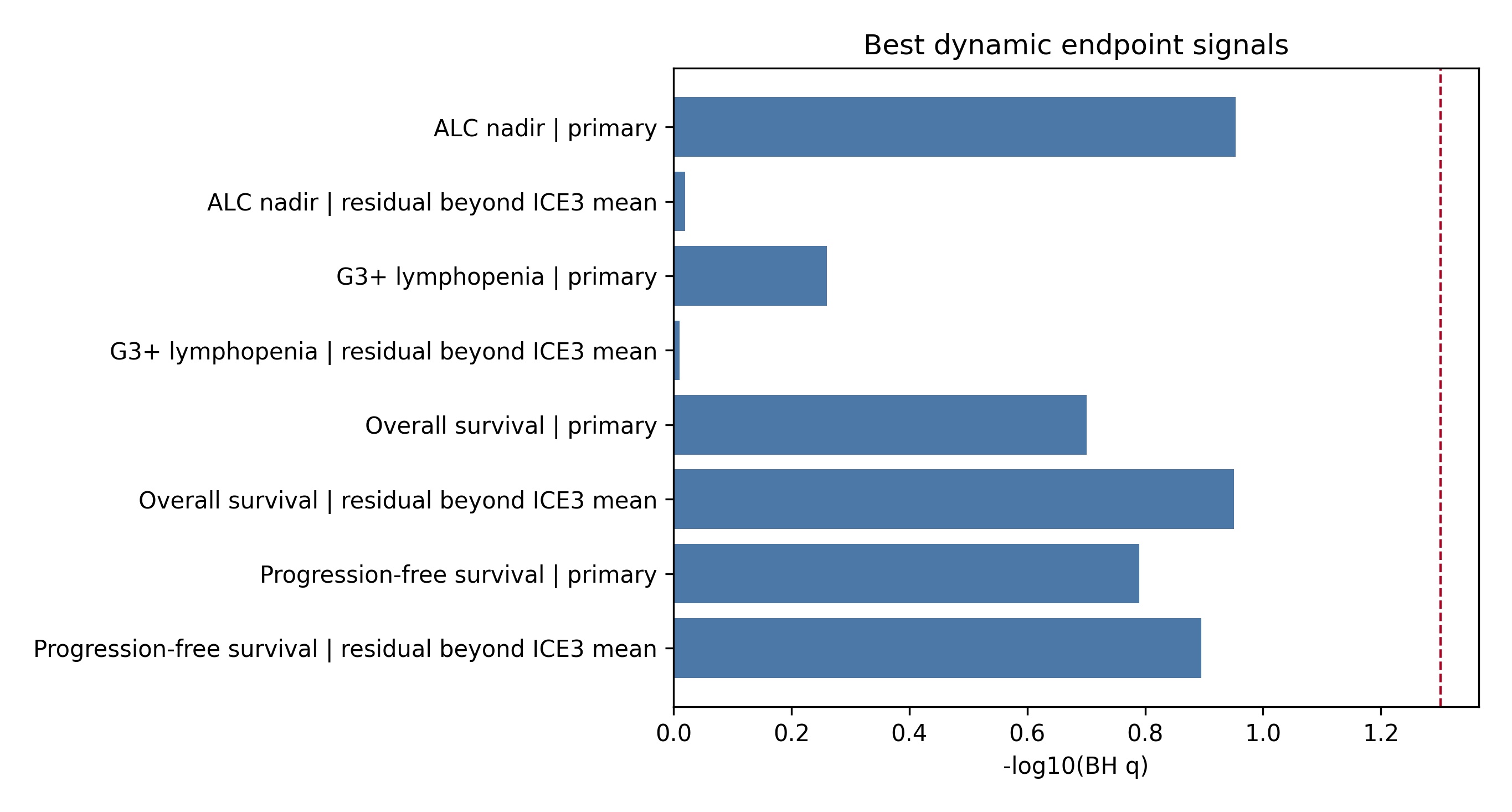

### Supplemental_Figure_S1_dynamic_endpoint_qvalues.png

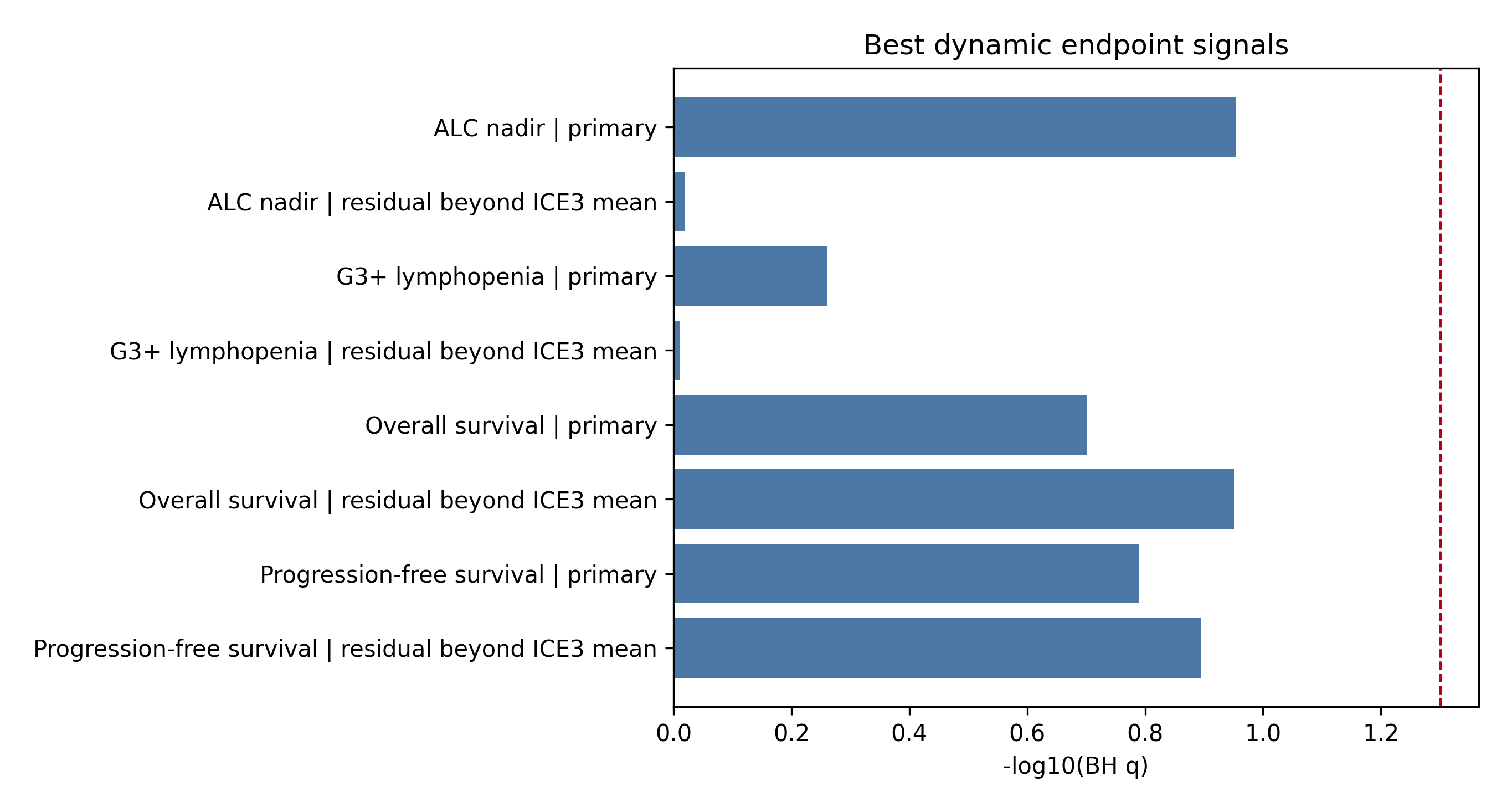

### Supplemental_Figure_S1_dynamic_endpoint_qvalues.tif

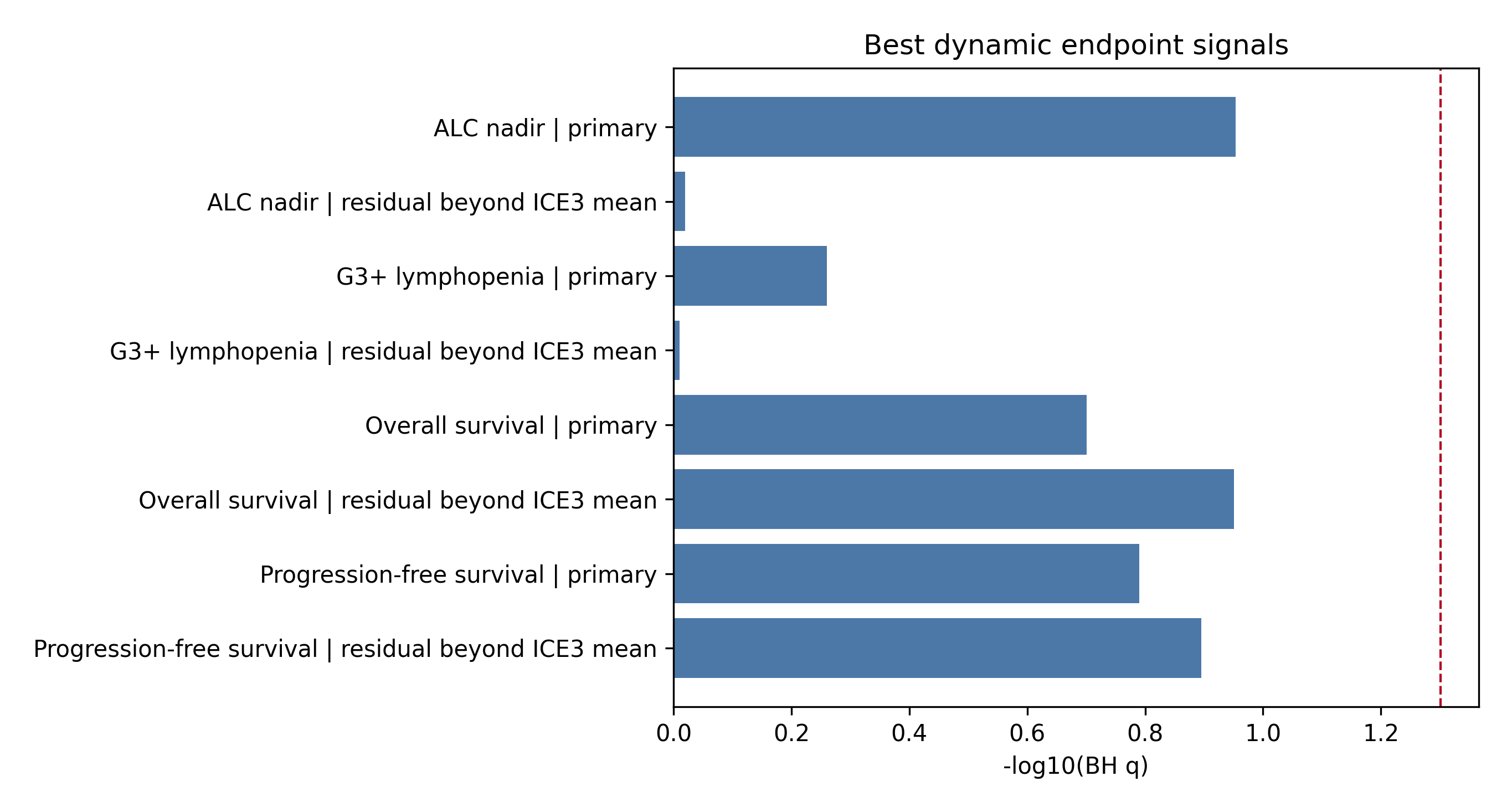

### Supplemental_Figure_S2_hedos_bdvhs.jpg

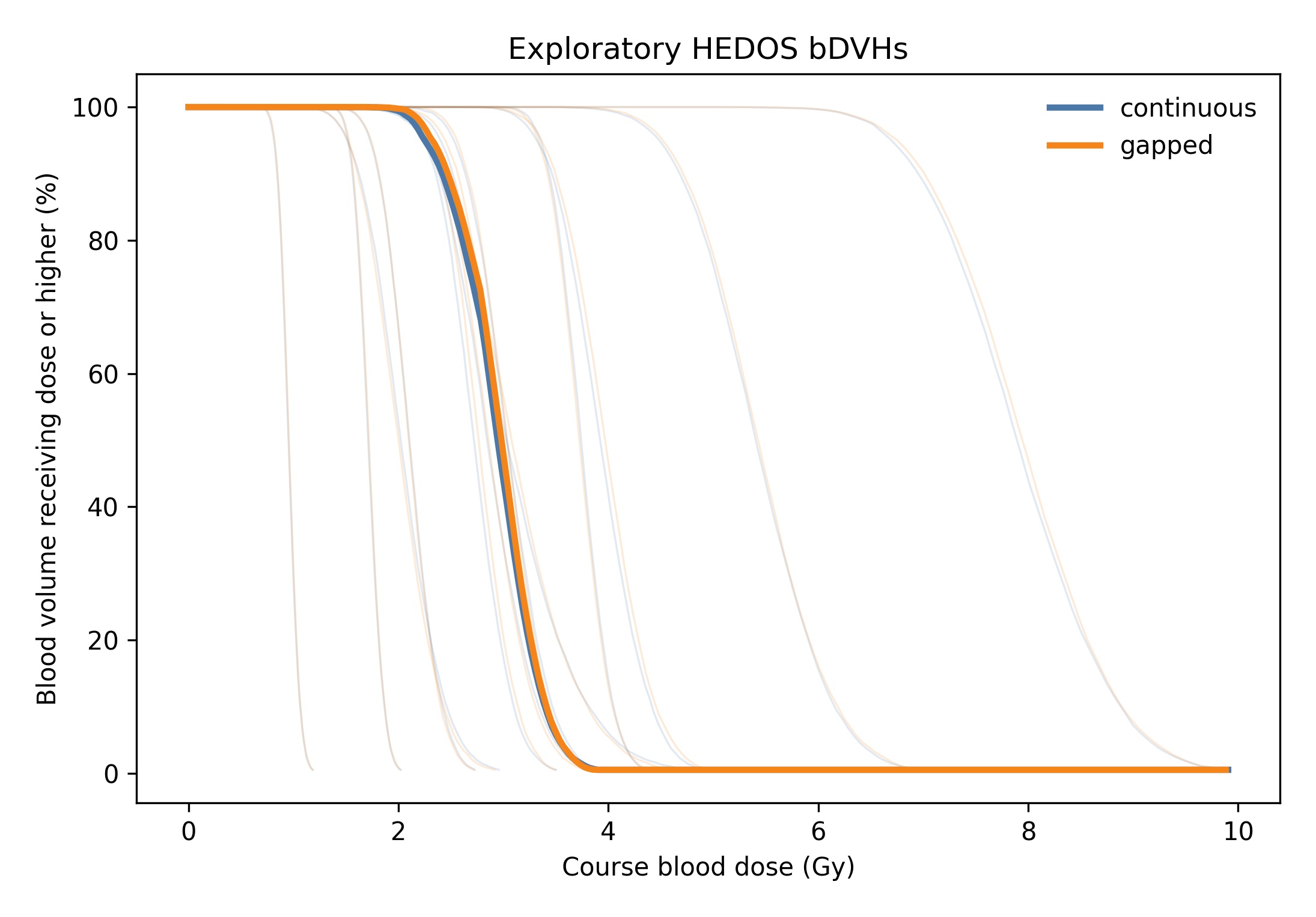

### Supplemental_Figure_S2_hedos_bdvhs.png

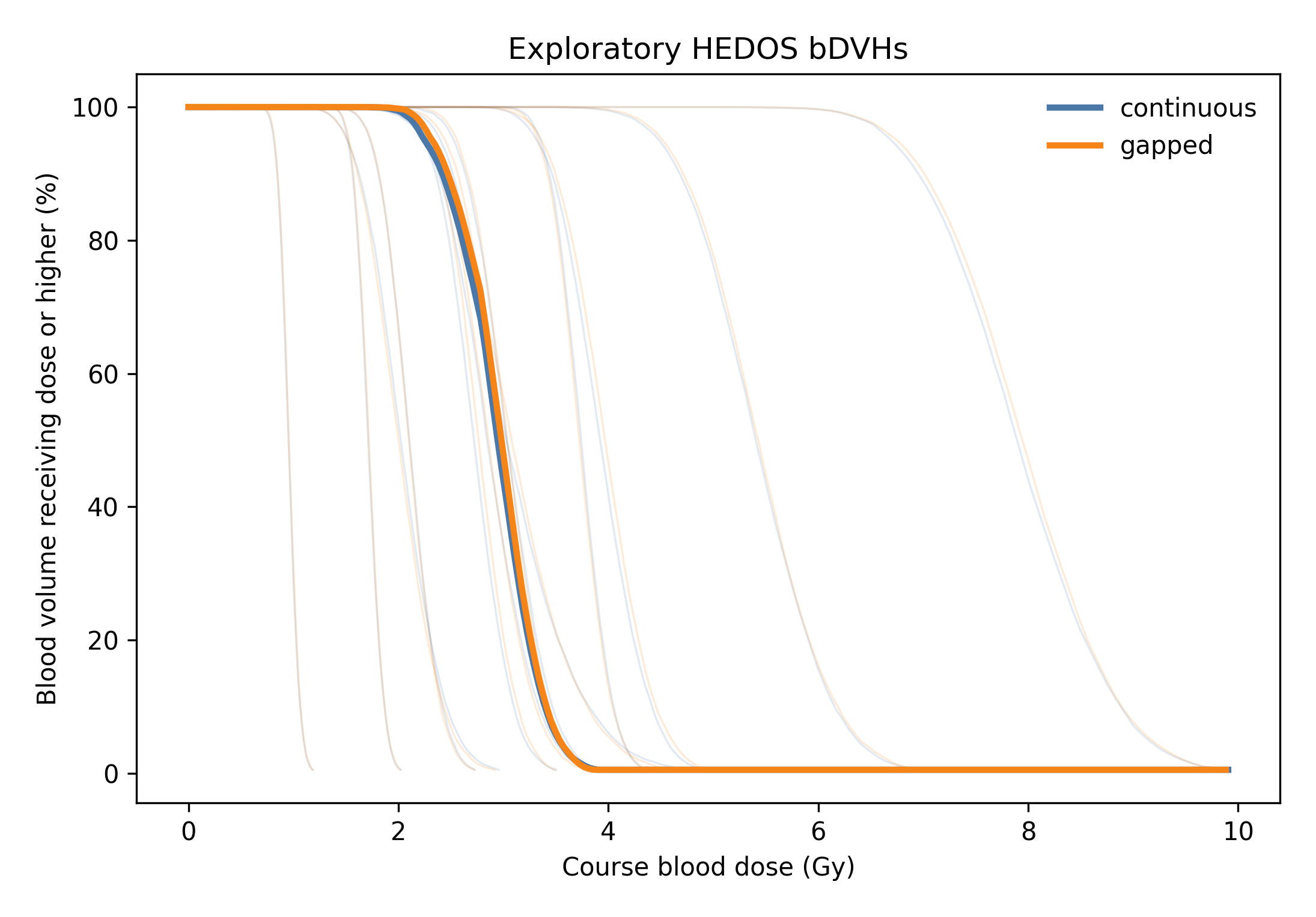

### Supplemental_Figure_S2_hedos_bdvhs.tif

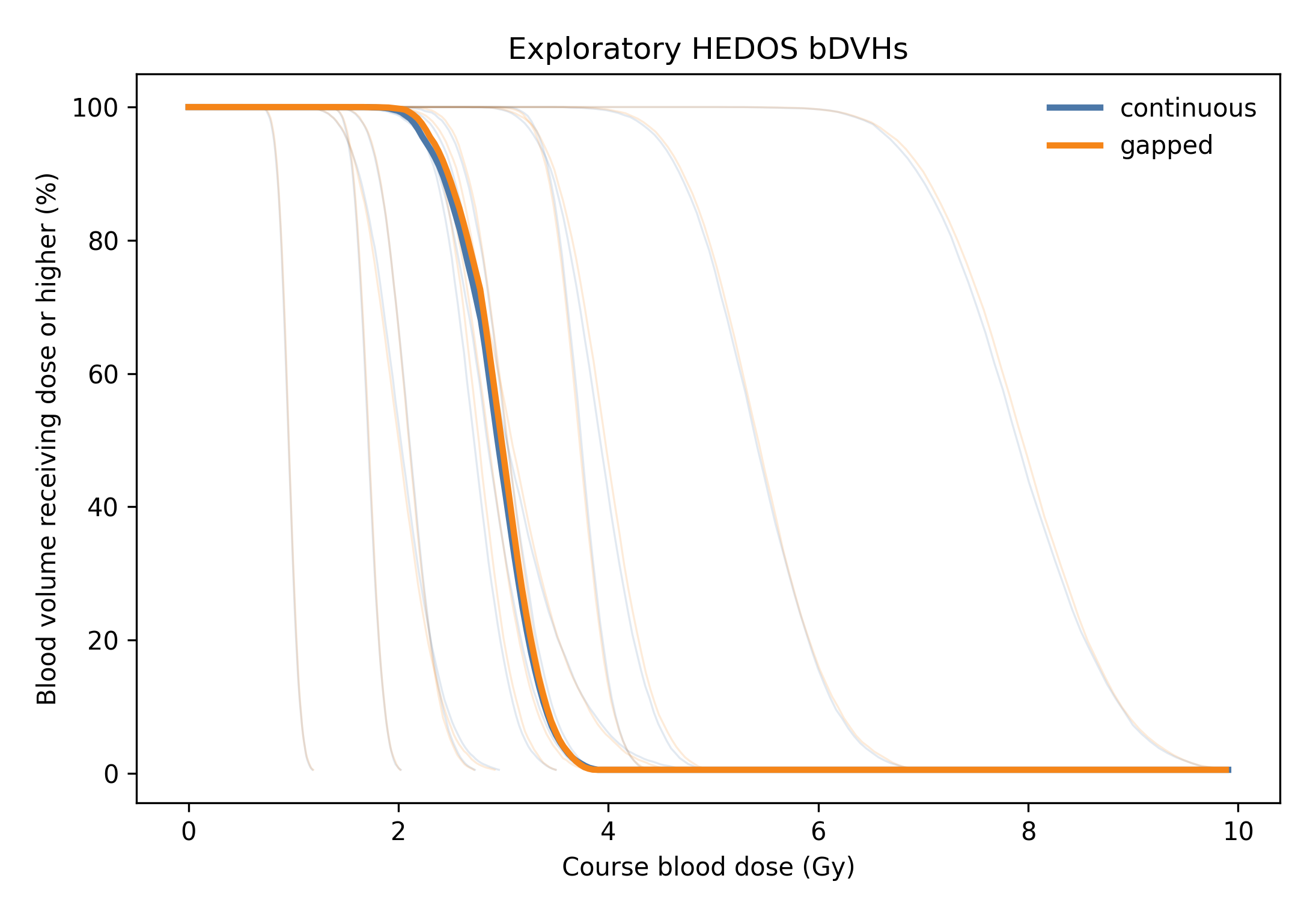

### Supplemental_Figure_S3_hedos_gap_delta.jpg

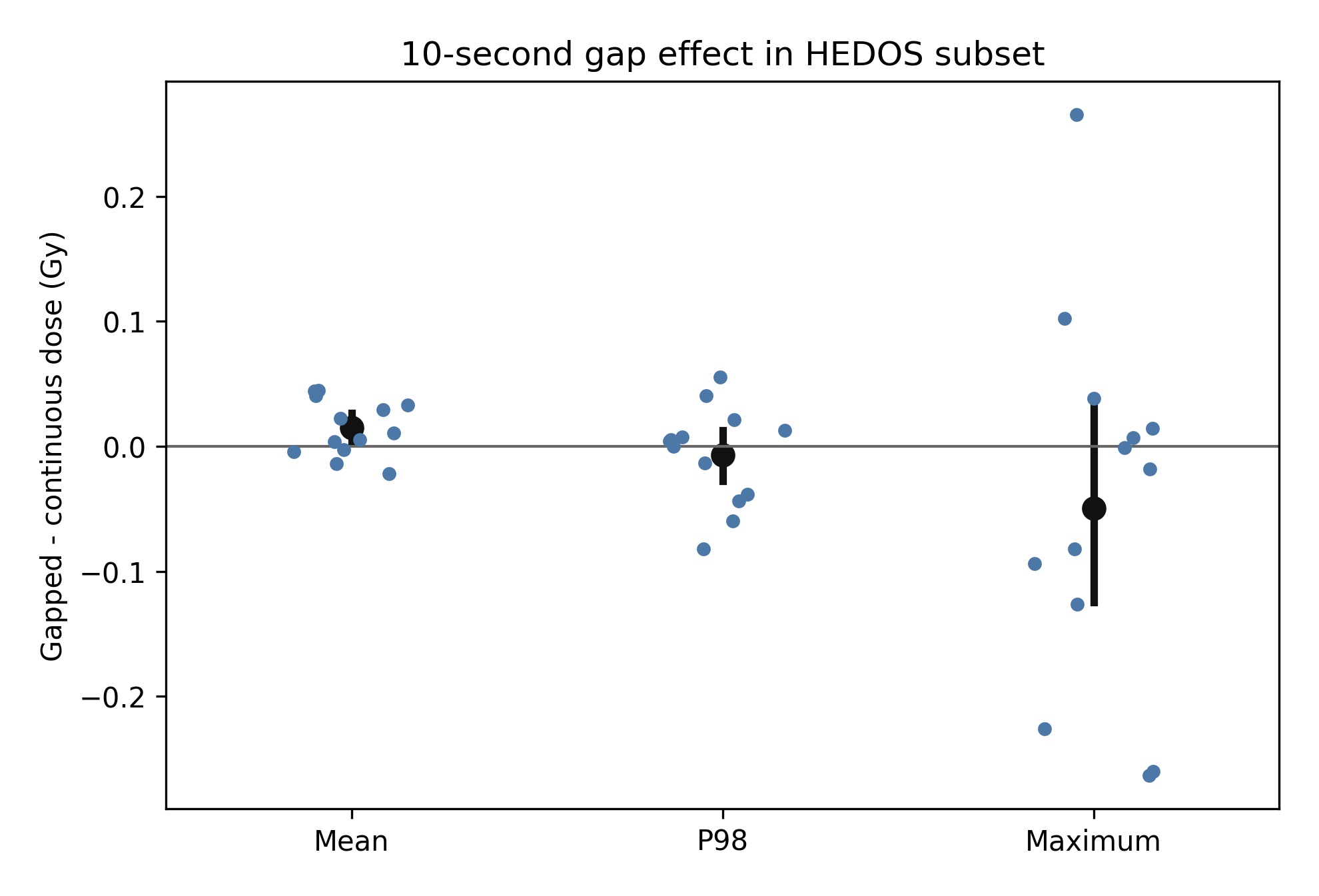

### Supplemental_Figure_S3_hedos_gap_delta.png

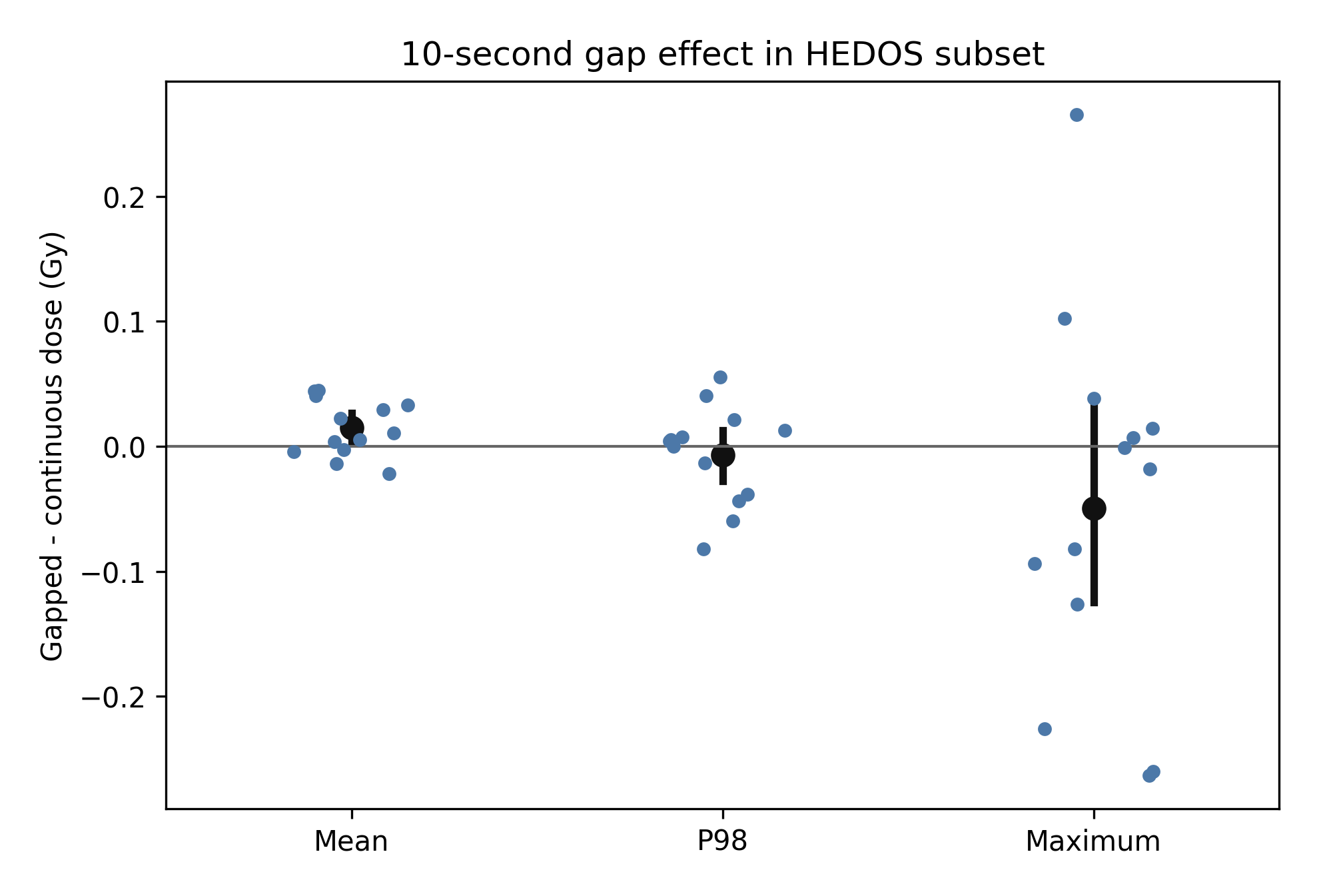

### Supplemental_Figure_S3_hedos_gap_delta.tif

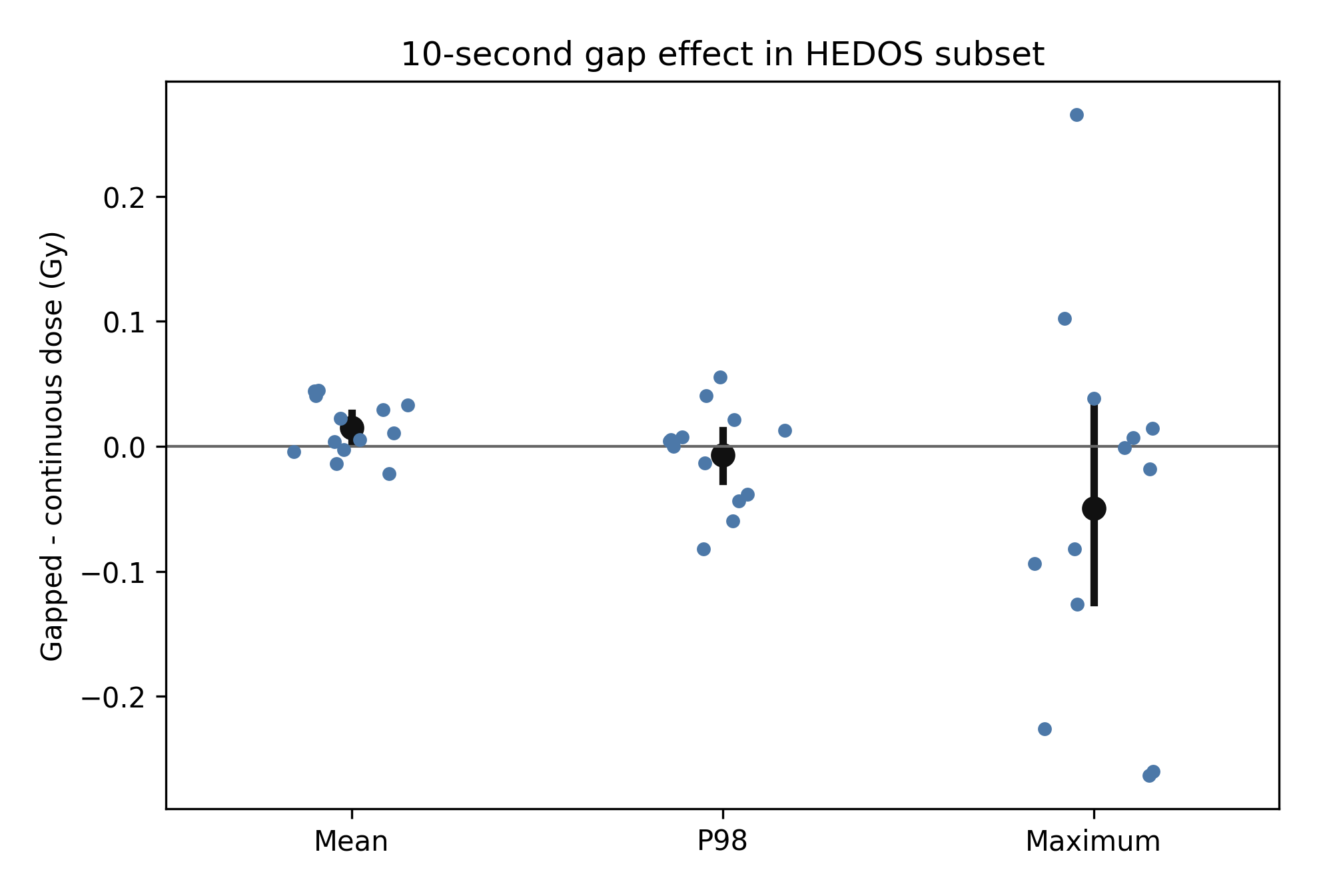

### Supplemental_Figure_S4_dynamic_correlations.jpg

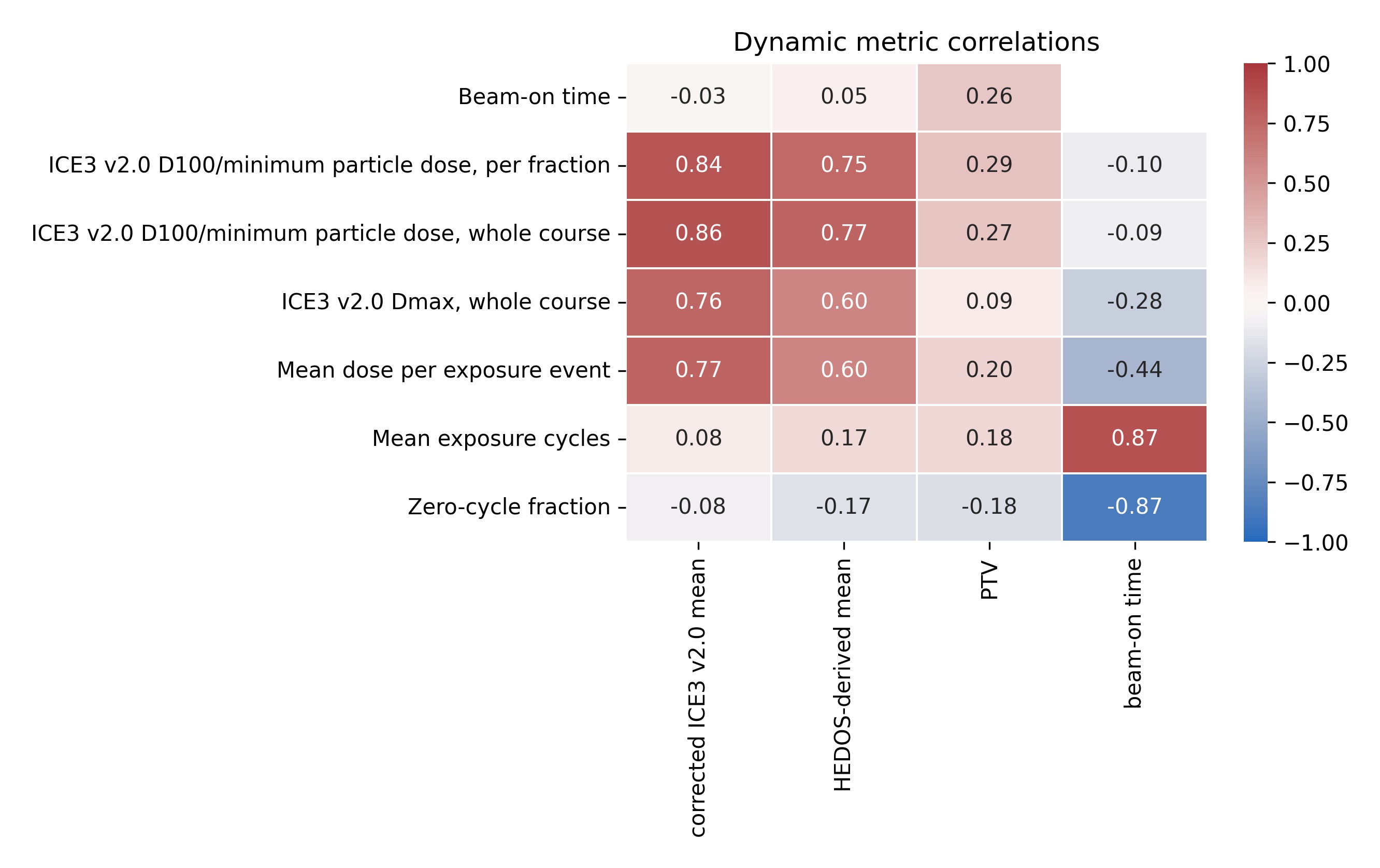

### Supplemental_Figure_S4_dynamic_correlations.png

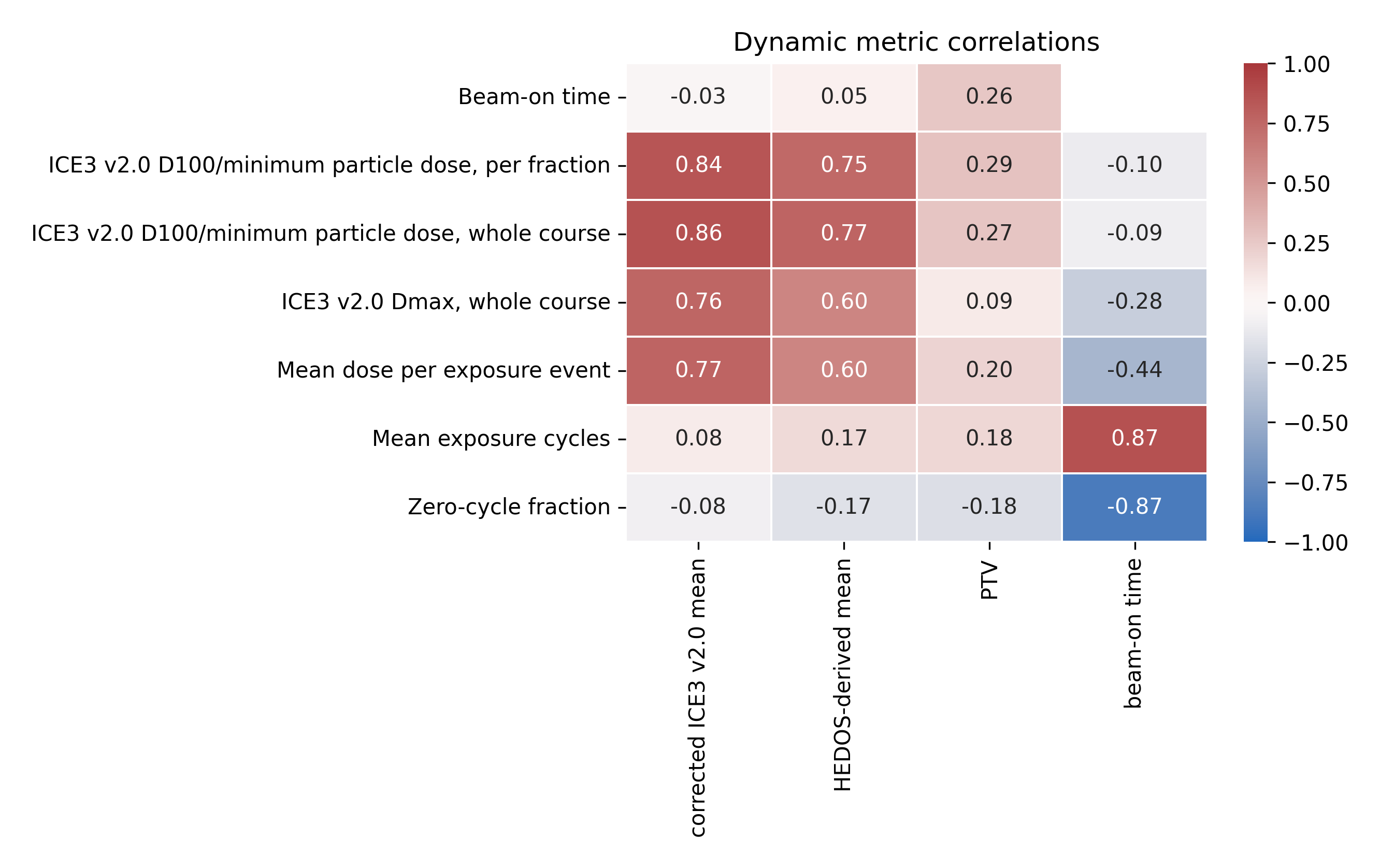

### Supplemental_Figure_S4_dynamic_correlations.tif

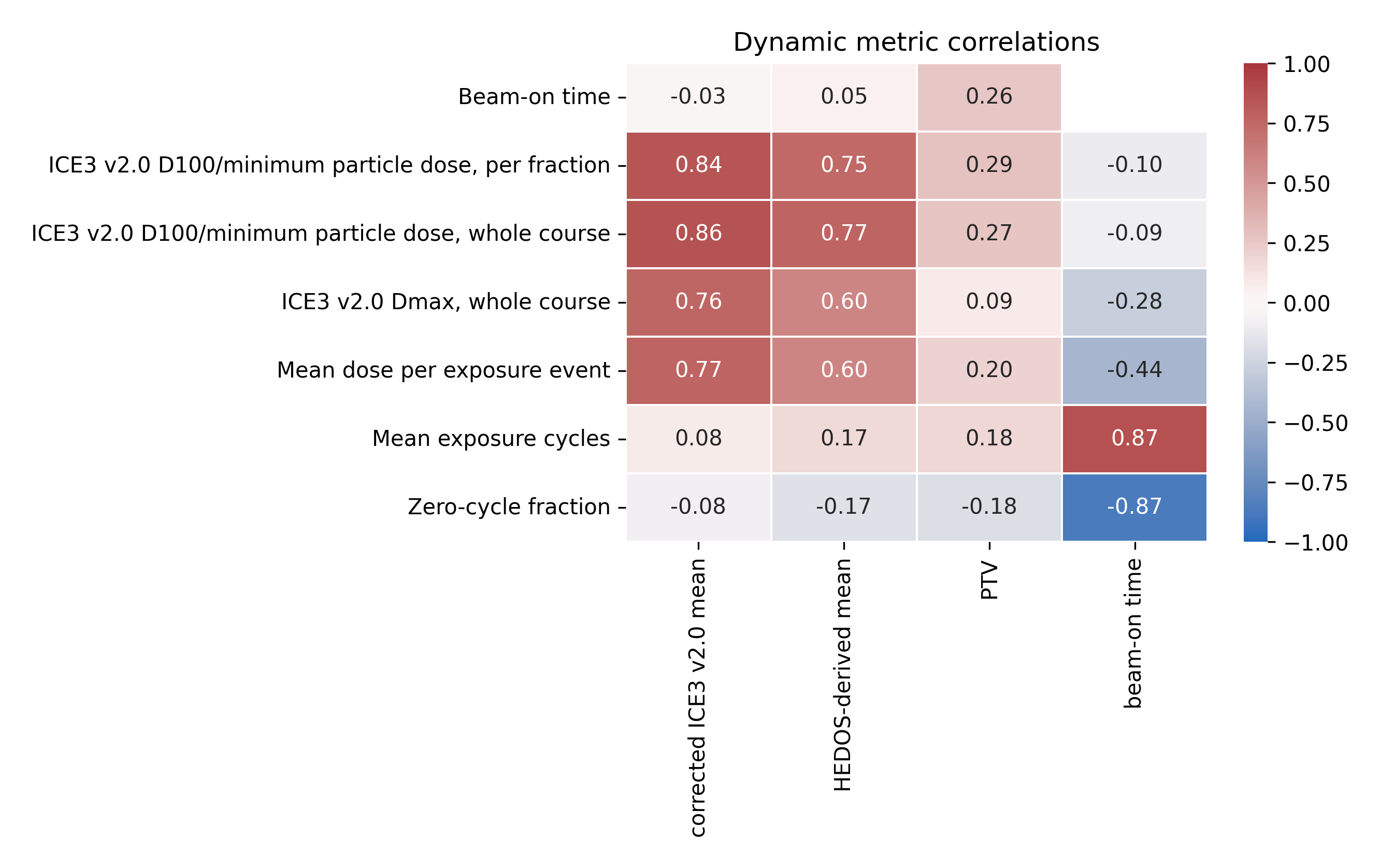
